# Distinct Biomarker Signatures of CMR-Defined Myocardial Remodeling in Moderate Aortic Stenosis

**DOI:** 10.64898/2026.10.26.26366467

**Authors:** Jan Wrobel, Jan Paul Janssen, Alina Beckmann, Jasmin Shamekhi, Mohammed Bahaaeldin, Julian Luetkens, Ibrahim Hawwari, Stephan Nienaber, Bernardo Franklin, Malte Kelm, Georg Nickenig, David Maintz, Stephan Baldus, Baravan al Kassou, Tobias Zeus, Lenhard Pennig, Victor Mauri

**Author notes:** Corresponding author: Priv.-Doz. Dr. med. Victor Mauri, Department of Cardiology, Heart Center Cologne, Faculty of Medicine and University Hospital, University of Cologne, Cologne, Germany, Kerpener Straße 62, 50937 Cologne, Germany. Drs. Wrobel and Janssen contributed equally.

## Abstract

**Background:** Moderate aortic stenosis (AS) is associated with adverse outcomes, yet the myocardial response to pressure overload varies substantially between patients and is incompletely reflected by valve hemodynamics. We aimed to determine whether circulating biomarkers of myocardial stress, injury, and fibrosis associate with cardiac magnetic resonance (CMR)-defined myocardial remodeling in moderate AS.

**Methods:** Eighty-nine patients with moderate AS prospectively underwent CMR (cine imaging, T1 mapping, extracellular volume fraction [ECV], late gadolinium enhancement [LGE]) with concurrent measurement of NT-proBNP, high-sensitivity troponin T, sST2, TIMP-1, and Galectin-3. Biomarker associations with the phenotype-defining CMR parameters were tested with adjustment for age, sex, and renal function and correction for multiple testing, and patients were additionally classified into 4 hierarchical remodeling phenotypes.

**Results:** Six of 20 biomarker-CMR associations remained significant after correction for multiple testing. NT-proBNP tracked LV mass index (beta=0.50), ECV (beta=0.35), and LGE burden (beta=0.43, all q<0.05). TIMP-1 and Galectin-3 were selectively associated with native T1 (both beta=0.35, q=0.012), and sST2 was inversely associated with LV mass index (beta=-0.27, q=0.046). TIMP-1 (adjusted OR 2.18 per SD, 95% CI 1.37-3.48, q=0.005) and NT-proBNP (OR 2.10, 1.19-3.69, q=0.026) increased with higher hierarchical remodeling phenotype. No biomarker was associated with echocardiographic mean gradient, peak velocity, or indexed valve area after correction.

**Conclusions:** Circulating biomarkers showed distinct associations with CMR measures of myocardial remodeling in moderate AS. Matrix-related biomarkers were associated with native T1, whereas NT-proBNP was associated across several remodeling domains. These signals were not measurably related to valve hemodynamics.

**Registration:** URL: https://clinicaltrials.gov/study/NCT05851209. Unique identifier: NCT05851209.

**Clinical Perspective What Is New?:**

- In patients with moderate aortic stenosis, myocardial remodeling on cardiac magnetic resonance was heterogeneous, and circulating biomarkers showed distinct associations with individual remodeling domains.
- The matrix-related biomarkers TIMP-1 and Galectin-3 were associated with native T1, whereas NT-proBNP was associated with several remodeling domains.

**What Are the Clinical Implications?:**

- No biomarker was associated with echocardiographic stenosis severity, suggesting that circulating biomarkers may add information on the myocardium that valve hemodynamics do not capture.
- If validated in larger cohorts, matrix-related biomarkers such as TIMP-1 could help select patients with moderate aortic stenosis for advanced myocardial tissue characterization.

## Introduction

Aortic stenosis (AS) is the most common valvular heart disease among older adults. Although assessment and intervention timing have traditionally centered on the severity of valvular obstruction, AS is increasingly recognized as a disease of both the valve and the myocardium, with the myocardial response to chronic pressure overload determining symptoms, outcomes, and recovery after aortic valve replacement (AVR) ^1–3^.

AS is associated with excess mortality and loss of life-years already at a moderate stage ^4^. Patients often remain below conventional thresholds for intervention, yet pressure overload may already drive extracellular matrix expansion, myocardial fibrosis, and subclinical ventricular dysfunction not captured by current standard clinical and echocardiographic evaluation. Consequently, patients with similar stenosis severity can exhibit markedly different myocardial responses, indicating that remodeling is a distinct dimension of disease only partly explained by valve obstruction and often unresolved by routine clinical assessment ^5,6^.

Cardiac magnetic resonance (CMR) is the reference standard for noninvasive myocardial tissue characterization. In AS, fibrosis evolves through both diffuse interstitial and replacement forms, yet mild-to-moderate AS remains underrepresented in CMR studies ^7,8^. Diffuse interstitial fibrosis can occur early in AS, is considered reversible and can regress after AVR ^9,10^, but is poorly detected by late gadolinium enhancement (LGE), a validated marker of the later, irreversible replacement fibrosis associated with adverse outcomes in AS ^11,12^. Native T1 mapping and extracellular volume fraction (ECV) address this limitation, allowing non-invasive assessment of diffuse myocardial remodeling ^13,14^.

In parallel, circulating biomarkers may reflect the biology underlying AS-driven remodeling. Natriuretic peptides and high-sensitivity troponins, markers of myocardial stress and injury, are associated with adverse remodeling and outcomes in AS ^15–17^, while markers of extracellular matrix turnover and fibrosis, including tissue inhibitor of metalloproteinase-1 (TIMP-1), Galectin-3, and soluble suppression of tumorigenicity 2 (sST2), have emerged as candidate indicators of pressure-overload cardiomyopathy ^18–24^. However, prior studies have largely related biomarkers to valve severity, symptoms, or outcomes rather than to specific myocardial remodeling phenotypes, and their relationship to CMR-defined tissue remodeling in moderate AS remains undefined.

The extent to which circulating biomarkers capture myocardial remodeling beyond conventional measures of valve hemodynamic severity in moderate AS remains uncertain. This study characterized myocardial remodeling by comprehensive CMR tissue phenotyping and examined its relationship with circulating markers of myocardial stress (sST2), injury (high-sensitivity troponin T), neurohormonal activation (NT-proBNP), and extracellular matrix turnover (TIMP-1 and Galectin-3). Patients were classified into imaging-defined phenotypes spanning LV hypertrophy, diffuse interstitial fibrosis, and focal replacement fibrosis. Associations between biomarkers and phenotype-defining CMR parameters, as well as, as an internal comparator, echocardiographic indices of stenosis severity, were evaluated using identical statistical models. Clinical follow-up was assessed on an explicitly exploratory basis.

## Methods

The data that support the findings of this study are available from the corresponding author upon reasonable request.

### Study Design and Patient Population

Adults with acquired, tricuspid moderate aortic stenosis (AS) were prospectively enrolled in the MODAS study (TRR259 project C06, ClinicalTrials.gov NCT05851209). Rationale and full eligibility criteria are provided in the Supplemental Methods. Between November 2023 and February 2026, 89 patients with complete baseline datasets were enrolled at the University Hospitals of Cologne and Bonn, Germany (Supplemental Figure S1). The study was approved by the institutional ethics committee (Cologne, 22-1458), complied with the Declaration of Helsinki, and all participants gave written informed consent.

### Cardiac Magnetic Resonance Imaging

Baseline CMR was performed on a 1.5-T system (Ingenia, Philips Healthcare, Best, the Netherlands) with cine imaging, native and post-contrast T1 mapping with ECV quantification, and late gadolinium enhancement (LGE). Acquisition, image analysis and reader blinding are detailed in the Supplemental Methods and Supplemental Table S1.

### CMR-Derived Remodeling Domains and Phenotype Classification

CMR measures were grouped a priori, independently of biomarker data, into three domains reflecting successive categories of the myocardial response to pressure overload: LV hypertrophy, diffuse interstitial fibrosis, and focal replacement fibrosis (constituent parameters in Figure 2). Each patient was also assigned to one of four mutually exclusive phenotypes of increasing severity: (1) no significant remodeling, (2) LV hypertrophy only (LV mass index >84 g/m2 in men, >69g/m2 in women) ^25^, (3) diffuse interstitial fibrosis (native T1 >985 ms or ECV >29%), and (4) LGE positive. Patients meeting more than one criterion were assigned the most advanced category, and those meeting none to phenotype 1. Thresholds were prespecified, derived from local healthy-volunteer reference values on the same system.

**Figure 1.**
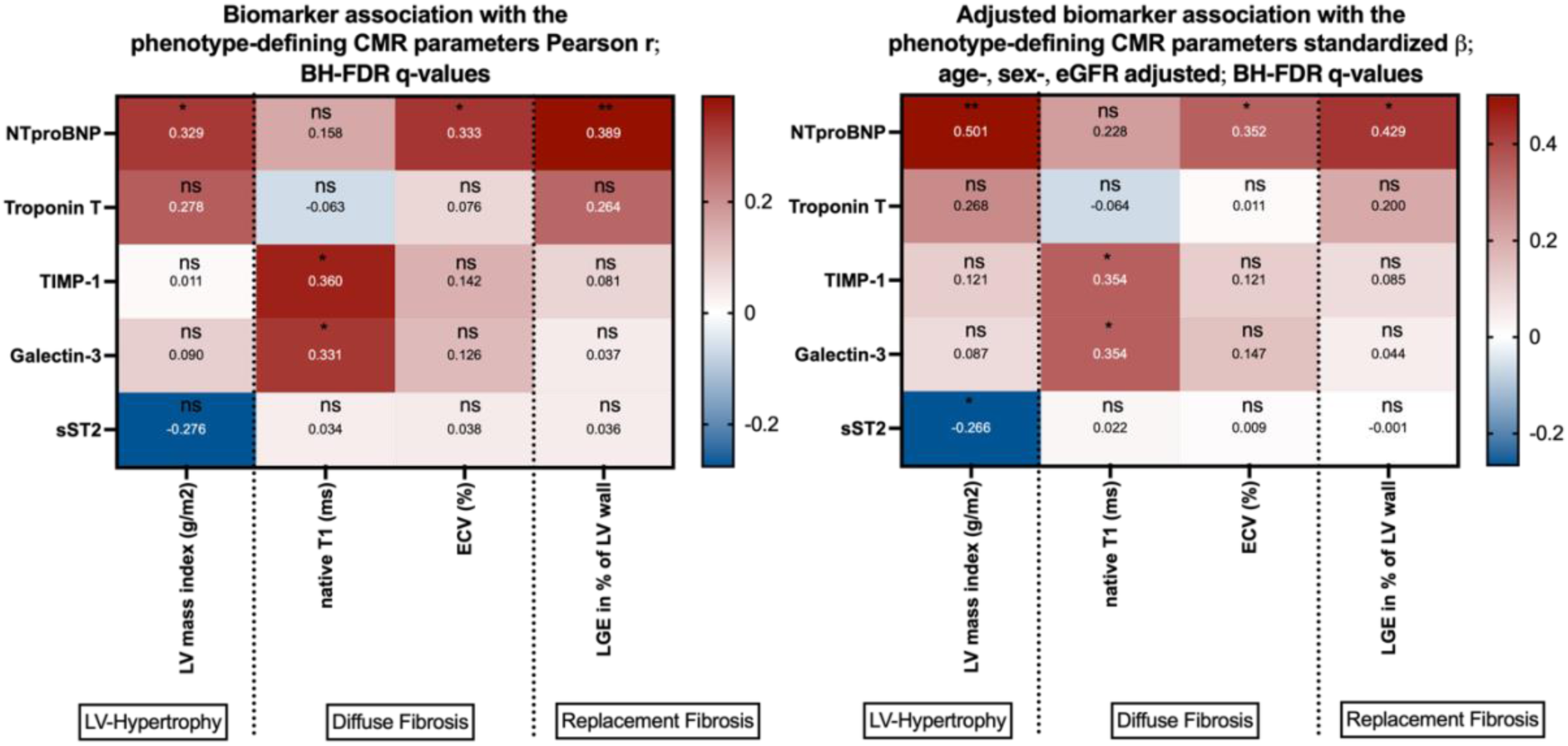
Association of log-transformed circulating biomarkers with the phenotype-defining CMR parameters in 89 patients with moderate AS. Two-panel figure showing associations of five log-transformed circulating biomarkers with LV mass index, native T1, ECV, and LGE burden; panel A shows univariable correlations and panel B estimates adjusted for age, sex, and eGFR.

**Figure 2.**
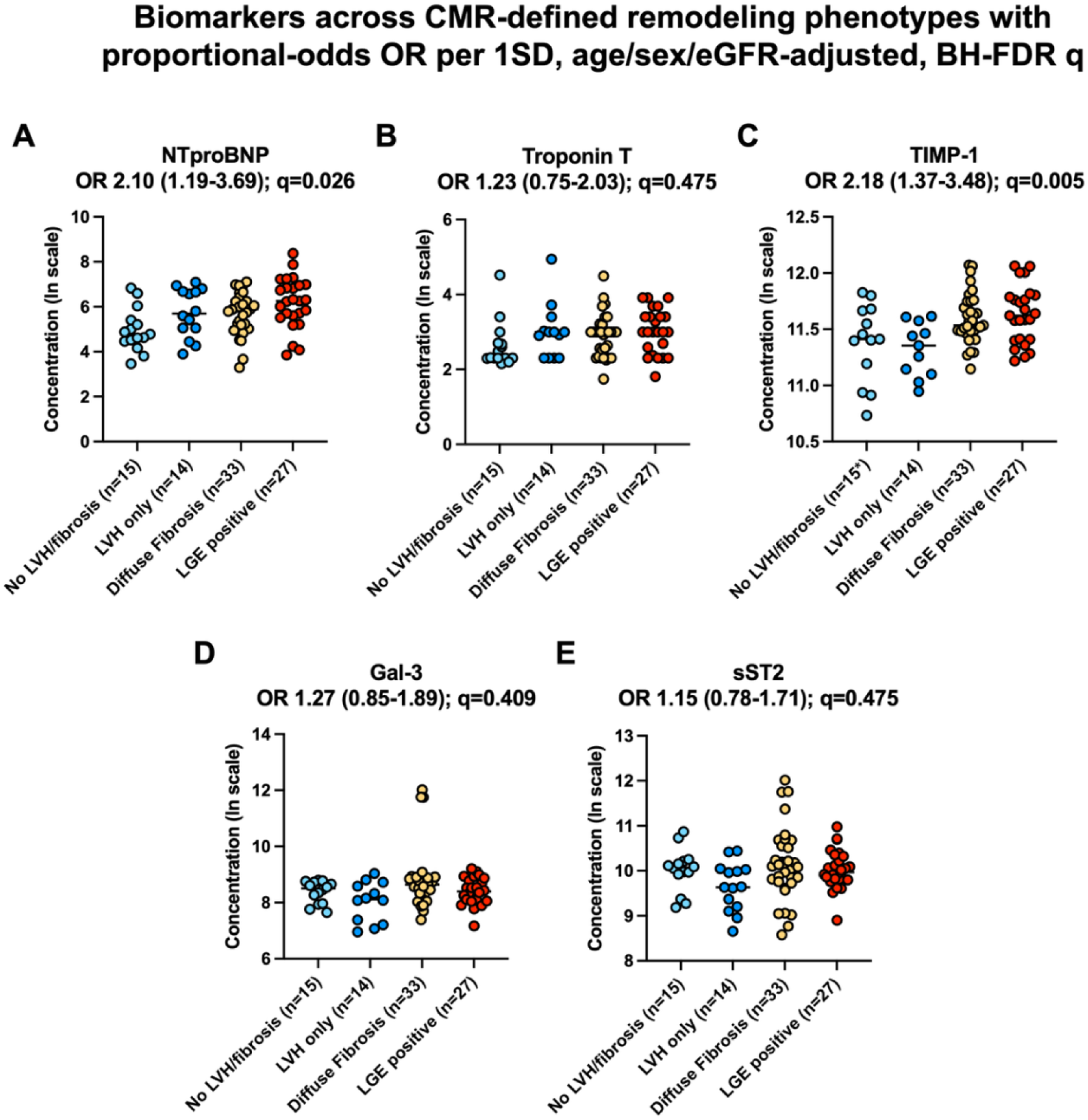
Biomarker concentrations (log scale) across CMR-defined remodeling phenotypes (no remodeling n=15, LV hypertrophy only n=14, diffuse interstitial fibrosis n=33, LGE positive n=27). Plots of the five biomarker concentrations on a logarithmic scale across the four CMR-defined remodeling phenotypes, with NT-proBNP and TIMP-1 higher in more advanced phenotypes.

### Circulating Biomarkers

Baseline EDTA plasma was stored at −80°C until batch analysis. sST2, TIMP-1 and Galectin-3 were measured by ELISA (R&D Systems/Bio-Techne, Minneapolis, Minnesota) and NT-proBNP and high-sensitivity troponin T by clinical immunoassay, all operators blinded to imaging and outcome data. Assay details are given in the Supplemental Methods.

### Composite Endpoint

The exploratory composite endpoint comprised aortic valve replacement (transcatheter or surgical) and echocardiographic progression to untreated, severe AS at follow-up (aortic valve area <1 cm², mean gradient >40 mmHg, or peak velocity >4 m/s). Time to event was measured from inclusion, and event-free patients were censored at last follow-up.

### Statistical Analysis

Continuous variables are summarized as median (Q1-Q3), and categorical variables as counts and percentages. Between-group comparisons were performed using the Mann-Whitney U test for continuous variables and the Fisher exact or chi-square test for categorical variables. Comparisons across the 4 remodeling phenotype categories used the Kruskal-Wallis or chi-square test.

The prespecified confirmatory analysis evaluated the associations between each log-transformed biomarker and the 4 phenotype-defining CMR parameters: LV mass index, native T1, extracellular volume fraction (ECV), and late gadolinium enhancement (LGE) burden.

Associations were assessed using linear regression models in which each biomarker was entered after adjustment for a prespecified covariate set of age, sex, and estimated glomerular filtration rate (eGFR), selected a priori as the principal noncardiac determinants of biomarker concentrations and of native T1 and ECV. The Benjamini-Hochberg procedure was used to control the false-discovery rate (FDR) across the 20 confirmatory tests, with q<0.05 considered statistically significant. Associations between biomarkers and all measured CMR parameters, including the phenotype-defining parameters, were considered exploratory and were FDR-controlled separately within that family of 55 tests (Supplemental Material). The covariate set was restricted to these three variables a priori, because the available sample size and event count do not support reliable estimation of models with a larger number of covariates. The association between biomarkers and remodeling phenotype category (1-4) was evaluated using proportional-odds ordinal logistic regression, with each standardized log-transformed biomarker modeled as a continuous predictor. Both unadjusted and adjusted models (age, sex, and eGFR) were examined, with FDR control applied across the 5 biomarkers.

Standardized regression coefficients (β) are reported, with 95% CIs for the corresponding unstandardized coefficients, and univariable Pearson correlation coefficients are presented alongside the adjusted estimates. LGE burden was coded as 0 in patients without LGE. For the exploratory clinical endpoint, effect estimates for continuous predictors are expressed per 1-SD increase, false-discovery-rate control was applied across the evaluated predictors, and event-free survival according to native T1 status was assessed by Kaplan-Meier analysis with the log-rank test.

As an internal control, analogous adjusted regression models and univariable Pearson correlations were used to evaluate associations between biomarkers and echocardiographic measures of aortic stenosis severity (mean transvalvular gradient, peak velocity, and indexed valve area). These 15 tests were FDR-controlled. Pearson correlations were also used to assess relationships between echocardiographic severity measures and the phenotype-defining CMR parameters, with FDR control across 12 tests.

Sensitivity analyses repeated the confirmatory biomarker-CMR analysis and ordinal regression models after exclusion of patients with infarct-pattern LGE.

Clinical follow-up analyses were prespecified as exploratory. Univariable Cox proportional hazards regression was used to examine associations of biomarkers, phenotype-defining CMR parameters, and indexed aortic valve area (iAVA) with the composite clinical endpoint. Elevated native T1 (>985 ms) was additionally examined by logistic regression. Effect-size expression, FDR handling, and Kaplan-Meier/log-rank details are provided in the Supplemental Statistical Methods. All tests were 2-sided. Statistical significance was defined as P<0.05, or as q<0.05 where FDR correction was applied. Statistical analyses were performed using IBM SPSS Statistics, version 28.0.2 (IBM Corp., NY, USA), and Python, version 3.14.6 (Python Software Foundation, OR, USA). Data visualization was performed using GraphPad Prism, version 11 (GraphPad Software, MA, USA).

## Results

### Baseline characteristics and outcomes

The cohort comprised 89 patients with moderate AS (median age 76.3 years (IQR 69.7-81.3), 22 (24.7%) women, Table 1). The median body mass index was 27.0 (24.0-31.0) kg/m2, with a median EuroSCORE II of 1.55 (0.90-2.91)%. Arterial hypertension (80.7%), hyperlipidemia (59.0%), and diabetes mellitus (34.9%) were common, and 12 (14.6%) patients had previous myocardial infarction and 27 (32.9%) atrial fibrillation. Median biomarker concentrations can be derived from Table 1 (additional characteristics in Supplemental Table S2).

**Table 1:**
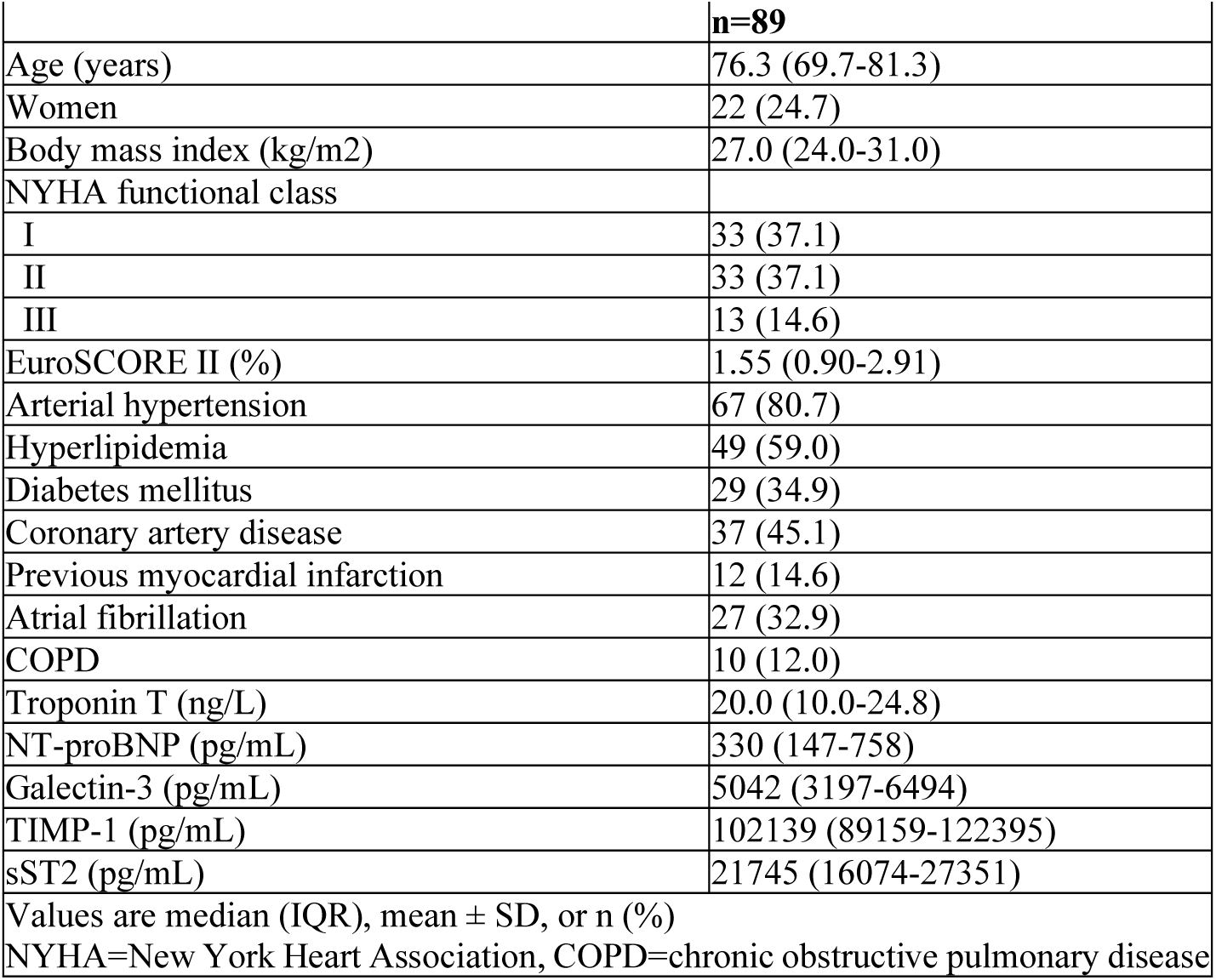
Patient-level characteristics.

| Table 1: Patient-level characteristics |  |
| --- | --- |
|  | **n=89** |
| Age (years) | 76.3 (69.7-81.3) |
| Women | 22 (24.7) |
| Body mass index (kg/m2) | 27.0 (24.0-31.0) |
| NYHA functional class |  |
| I | 33 (37.1) |
| II | 33 (37.1) |
| III | 13 (14.6) |
| EuroSCORE II (%) | 1.55 (0.90-2.91) |
| Arterial hypertension | 67 (80.7) |
| Hyperlipidemia | 49 (59.0) |
| Diabetes mellitus | 29 (34.9) |
| Coronary artery disease | 37 (45.1) |
| Previous myocardial infarction | 12 (14.6) |
| Atrial fibrillation | 27 (32.9) |
| COPD | 10 (12.0) |
| Troponin T (ng/L) | 20.0 (10.0-24.8) |
| NT-proBNP (pg/mL) | 330 (147-758) |
| Galectin-3 (pg/mL) | 5042 (3197-6494) |
| TIMP-1 (pg/mL) | 102139 (89159-122395) |
| sST2 (pg/mL) | 21745 (16074-27351) |
| Values are median (IQR), mean ± SD, or n (%) |  |
| NYHA=New York Heart Association, COPD=chronic obstructive pulmonary disease |  |

### Imaging characteristics

The median mean aortic gradient was 17.0 (13.7-22.0) mmHg, peak velocity 2.80 (2.50-3.10) m/s, and indexed AVA 0.69 (0.59-0.78) cm2/m2, confirming moderate-range hemodynamics (Table 2). On CMR, the median LV mass index was 82.9 (73.2-93.9) g/m2, maximum LV wall thickness 12.6 (11.4-14.1) mm, native T1 994 (973-1027) ms, ECV 25.9 (24.0-28.6)%, and LVEF 69 (62-75)%. Native T1 was elevated (>985 ms) in 52 patients (58.4%), ECV exceeded 29% in 20 of 86 (23.3%), and LGE was present in 27 patients (30.3%), of whom 12 (44.4%) showed an infarct pattern (Table 2).

**Table 2:** Imaging characteristics and outcome.

| **Table 2: Imaging characteristics and outcome** |  |
| --- | --- |
|  | **n=89** |
| Peak AV gradient (mmHg) | 31.9 (24.0-39.0) |
| Mean AV gradient (mmHg) | 17.0 (13.7-22.0) |
| Peak AV velocity (m/s) | 2.80 (2.50-3.10) |
| Indexed AVA (cm<sup>2</sup>/m<sup>2</sup>) | 0.69 (0.59-0.78) |
| Moderate/severe mitral regurgitation | 10 (11.5) |
| Moderate/severe tricuspid regurgitation | 14 (16.3) |
| Cell volume (mL) | 113.9 (97.5-131.3) |
| LV mass (g) | 162.9 (136.9-189.7) |
| LV mass index (g/m<sup>2</sup>) | 82.9 (73.2-93.9) |
| Max LV wall thickness (mm) | 12.6 (11.4-14.1) |
| ECV fraction (%) | 25.9 (24.0-28.6) |
| iECV (mL/m<sup>2</sup>) | 21.0 (16.9-24.9) |
| Native T1 overall (ms) | 994 (973-1027) |
| Native T1 basal (ms) | 1004 (982-1032) |
| Native T1 >985 ms | 52 (58.4) |
| LGE positive | 27 (30.3) |
| Infarct-pattern LGE | 12 (13.5) |
| Non-infarct LGE | 15 (16.9) |
| LGE (% of LV mass, 0 if LGE-negative) | 0.0 (0.0-1.3) |
| LVEF (%) | 69 (62-75) |
| Median follow-up (days) | 362 (251-422) |
| Death (any, descriptive) | 1 (1.1) |
| Composite events (AVR or severe-AS progression*) | 18 (20.2) |
| Values are median (IQR), mean ± SD, or n (%) | |
| AV=aortic valve, AVR=aortic valve replacement. |  |
| * Progression to severe aortic stenosis AVA <1cm<sup>2</sup>, or mean aortic pressure gradient >40mmHg, or peak aortic velocity >4m/s at follow-up |  |

### Association of circulating biomarkers with CMR-defined remodeling

Six biomarker-CMR associations remained significant after false discovery rate (FDR) correction, with distinct patterns across the remodeling domains (Figure 1, full coefficients in Supplemental Table S4). NT-proBNP showed the broadest pattern, associating with LV mass index (beta=0.50, q=0.004), ECV (beta=0.35, q=0.046), and LGE burden (beta=0.43, q=0.012). The matrix turnover markers were domain-selective: TIMP-1 and Galectin-3 each associated with native T1 (both beta=0.35, q=0.012) and with no other parameter. sST2 was inversely associated with LV mass index (beta=-0.27, q=0.046). No troponin T association survived correction. Univariable and adjusted estimates were concordant (Figure 1A vs 1B). In the exploratory extension to all measured CMR parameters (55 tests), the same pattern held, with additional NT-proBNP associations with iECV, matrix volume, LV mass, and LVEF and a Galectin-3 association with basal native T1 surviving FDR (Supplemental Figure S4, Supplemental Table S4). No further troponin T or sST2 association survived.

### Biomarker profiles across remodeling phenotypes

Patients were distributed across the four prespecified remodeling phenotypes as 15 (no remodeling), 14 (LV hypertrophy only), 33 (diffuse interstitial fibrosis), and 27 (LGE positive) (Table 3). In proportional-odds models, TIMP-1 was associated with higher remodeling phenotype in both unadjusted and adjusted analyses (unadjusted OR 2.11 per SD, 95% CI 1.37– 3.23, q=0.003, adjusted OR 2.18, 1.37–3.48, q=0.005), as was NT-proBNP (unadjusted OR 2.03, 1.32–3.13, q=0.003, adjusted OR 2.10, 1.19–3.69, q=0.026), with estimates strengthening rather than attenuating after adjustment. Troponin T, Galectin-3, and sST2 were not associated with remodeling phenotype (all q>=0.39) (Figure 2). Descriptive Kruskal-Wallis comparisons were concordant (NT-proBNP p=0.008, TIMP-1 p=0.005). Age, BMI, NYHA class, EuroSCORE II, sex distribution, and all echocardiographic severity measures were comparable across phenotypes (all p>0.05, Table 3), as were event rates.

**Table 3:** CMR-based remodeling phenotypes.

| **Table 3: CMR-based remodeling phenotypes** |  |  |  |  |  |
| --- | --- | --- | --- | --- | --- |
|  | **Category I**<br>**No LVH/**<br>**fibrosis**<br>**(n=15)** | **Category II**<br>**LVH only**<br>**(n=14)** | **Category III**<br>**Diffuse Fibrosis**<br>**+/- LVH**<br>**(n=33)** | **Category IV:**<br>**LGE positive +/-**<br>**diffuse fibrosis**<br>**(n=27)** | **p-value** |
| Age (years) | 70.1 (66.3-75.0) | 75.3 (62.6-85.1) | 76.8 (70.4-81.7) | 78.4 (75.1-81.3) | 0.053 |
| Women, n (%) | 2 (13%) | 5 (36%) | 10 (30%) | 5 (19%) | 0.3817 |
| Body mass index<br>(kg/m<sup>2</sup>) | 26.0 (23.8-29.0) | 25.5 (24.6-30.2) | 28.0 (23.0-31.0) | 28.0 (25.0-31.0) | 0.6045 |
| **Clinical factors** |  |  |  |  |  |
| NYHA functional class |  |  |  |  | 0.521 |
| I | 6 (40.0) | 8 (57.1) | 15 (45.4) | 9 (33.3) |  |
| II | 7 (46.7) | 2 (14.3) | 13 (39.4) | 16 (59.3) |  |
| III | 2 (13.3) | 4 (28.6) | 5 (15.2) | 2 (7.4) |  |
| EuroSCORE II (%) | 1.48 (0.84-1.90) | 1.12 (0.69-2.58) | 1.51 (1.08-2.70) | 2.09 (1.25-2.99) | 0.2116 |
| **Cardiac biomarkers** |  |  |  |  |  |
| Troponin (ng/L) | 10.1 (10.0-14.7) | 19.3 (10.0-20.4) | 20.0 (10.4-20.0) | 20.0 (12.7-30.0) | 0.0889 |
| NT-proBNP (pg/mL) | 119 (90-202) | 300 (157-793) | 360 (183-512) | 521 (272-1037) | 0.0075 |
| Galectin-3 (pg/mL) | 4922 (3134-6078) | 3361 (1546-5556) | 5402 (4027-6873) | 4423 (3258-6616) | 0.2084 |
| TIMP-1 (pg/mL) | 89940 (60257-113191) | 85360 (67648-100666) | 102139 (96926-126457) | 111397 (90130-131478) | 0.005 |
| sST2 (pg/mL) | 23685 (20646-26786) | 15320 (10370-22385) | 23944 (17265-27821) | 21368 (18267-29677) | 0.1637 |
| **Echocardiographic measures at baseline** |  |  |  |  |  |
| Mean aortic valve<br>gradient (mmHg) | 15.0 (11.3-18.5) | 19.0 (16.0-26.0) | 18.9 (13.8-23.3) | 16.0 (14.0-21.2) | 0.1847 |
| Indexed aortic valve<br>area (cm<sup>2</sup>/m<sup>2</sup>) | 0.76 (0.71-0.84) | 0.67 (0.58-0.73) | 0.67 (0.58-0.77) | 0.68 (0.58-0.77) | 0.0837 |
| **Outcome at 1 year** |  |  |  |  |  |
| Median follow-up<br>(days) | 365 (356-419) | 330 (200-388) | 344 (223-411) | 370 (255-484) | 0.4382 |
| AVR or AS<br>progression* | 2 (13.3) | 1 (7.1) | 10 (30.3) | 5 (18.5) | 0.2558 |
| All-cause Death | 0 (0.0) | 0 (0.0) | 0 (0.0) | 1 (3.7) | 0.629 |
| Values are median (IQR), mean ± SD, or n (%)<br>AVR=aortic valve replacement, LVH=Left ventricular hypertrophy, NYHA=New York Heart Association<br>* Progression to severe aortic stenosis AVA <1cm<sup>2</sup>, or mean aortic pressure gradient >40mmHg, or peak aortic velocity >4m/s at follow-up | | | | | |

### Biomarkers and valve hemodynamics

None of the circulating biomarkers was associated with echocardiographic measures of aortic valve hemodynamic severity in either statistical specification (Pearson minimum p=0.062, adjusted minimum p=0.080, all q>=0.47, Figure 3A). The same severity indices partly associated with parameters of myocardial remodeling: mean gradient correlated with LV mass index (r=0.31, q=0.0498), with concordant nonsignificant trends for peak velocity and for indexed valve area with native T1, and no severity index related to ECV or LGE burden (Figure 3B). Given the restricted range of valve hemodynamics, these null associations should be interpreted as absence of a detectable relationship in this cohort, not as proof of statistical independence.

**Figure 3.**
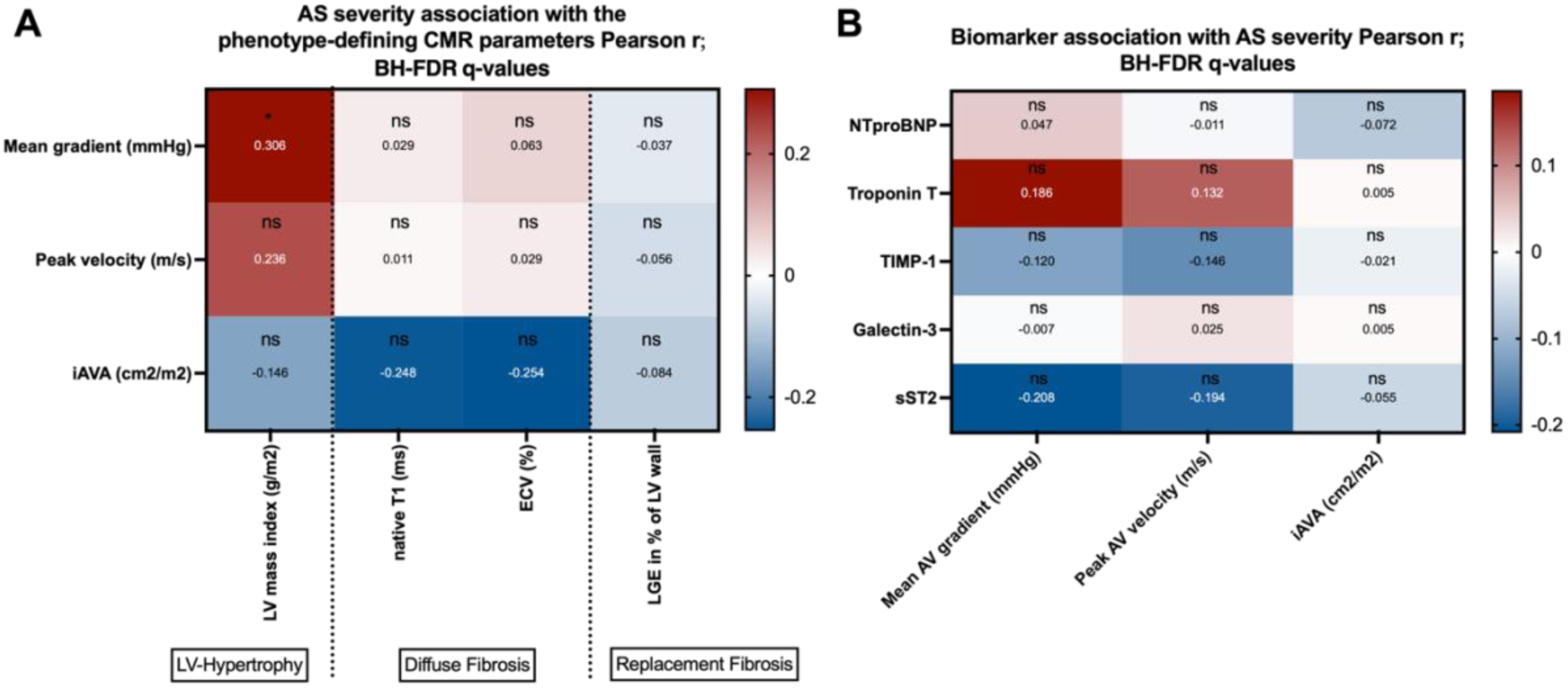
Associations between phenotype-defining CMR parameters, biomarkers and indices of AS severity. Panel A shows associations of the five biomarkers with mean gradient, peak velocity, and indexed aortic valve area; panel B shows correlations of these severity indices with the phenotype-defining CMR parameters.

### Sensitivity analyses

Excluding the 12 patients with infarct-pattern LGE (n=77), effect magnitudes were essentially unchanged, although two associations no longer met the FDR-threshold in the reduced sample. NT-proBNP remained associated with LV mass index (β=0.53, q=0.006) and Galectin-3 with native T1 (β=0.37, q=0.026), whereas the TIMP-1–native T1 association, similar in magnitude to that in the full cohort (β=0.33 vs 0.35), no longer survived correction (p=0.010, q=0.050), as was the case for sST2–LV mass index (β=−0.31, p=0.009, q=0.050). The TIMP-1 association with remodeling phenotype category persisted (ordinal OR 2.06, 95% CI 1.24–3.42, p=0.005) (Supplemental Table S5).

### Exploratory analysis of clinical follow-up

Over a median follow-up of 362 days (IQR 251-422), 18 patients (20.2%) reached the exploratory composite endpoint (15 aortic valve replacements and 3 untreated progressions to severe AS as first events). One patient died during follow-up. Patients reaching the endpoint more frequently had previous myocardial infarction (35.3% vs 9.2%) and less frequently hyperlipidemia (33.3% vs 66.2%), with no differences in other baseline, laboratory, or imaging characteristics apart from a trend toward smaller indexed valve area (0.59 vs 0.70 cm2/m2, p=0.073) (Supplemental Tables S6 and S7). The incidence of AVR or progression to severe AS did not differ across the CMR-defined remodeling categories (13.3% in Category I, 7.1% in Category II, 30.3% in Category III, and 18.5% in Category IV, p=0.256) (Table 3). Similarly, all-cause mortality was low, with one death occurring in Category IV (p=0.629).

In exploratory analyses of the 18-event composite, center-specific native T1 thresholds (>985 ms) showed a nonsignificant trend (OR 3.04, 95% CI 0.91–10.14, p=0.071, log-rank p=0.185, Cox HR 2.10, 95% CI 0.69–6.41, p=0.194). Native T1 modeled continuously was not associated with the endpoint (OR 1.36 per SD, 95% CI 0.82–2.25, p=0.235, Cox HR 1.08, 95% CI 0.73–1.59, p=0.713), and no threshold between 955 and 1030 ms reached significance (Supplemental Figure S3). Given the limited number of events, this cohort was not powered for prognostic inference, and no prognostic conclusions are drawn.

## Discussion

In this prospective study of 89 patients with moderate AS undergoing comprehensive CMR tissue characterization and concurrent biomarker profiling, the principal findings are threefold. First, myocardial remodeling in moderate AS is heterogeneous and only partly explained by the severity of valvular obstruction. Second, circulating biomarkers demonstrate domain-specific associations with CMR-defined remodeling: markers of extracellular matrix turnover are selectively associated with diffuse interstitial fibrosis and increase across hierarchical remodeling phenotypes, whereas NT-proBNP is associated with remodeling across multiple domains. Third, none of the five biomarkers was associated with any echocardiographic index of stenosis severity, indicating that the myocardial and valvular axes of the disease are dissociable and separately measurable within a single cohort.

### Relationship between biomarkers and valve hemodynamics

Assessment and intervention timing in AS have traditionally centered on the severity of valvular obstruction and on the assumption that the ventricular response parallels hemodynamic load. In this cohort, mean gradient was associated with LV mass index, whereas biomarker concentrations were not detectably associated with the three echocardiographic severity indices. Substantial interindividual variation in myocardial remodeling at similar stenosis grades has also been described in prior CMR studies ^7,8,26^. However, the narrow hemodynamic range imposed by the inclusion criteria attenuates correlations and limits formal comparison with associations involving CMR measures. Accordingly, the present data suggest, but do not prove, that circulating biomarkers provide information complementary to routine valve hemodynamics. Consistent with the cohort’s position on the disease continuum, native T1 and iECV were lower than values reported in severe AS ^27,28^, in keeping with progressive accumulation of diffuse fibrosis across the disease course, although differences in scanner platform and sequence cannot be excluded.

### Domain-specific biomarker signatures

Six of 20 prespecified biomarker-CMR associations remained significant after false-discovery-rate correction, and their distribution was not uniform. NT-proBNP demonstrated the broadest pattern, with associations across LV mass index, ECV, and LGE burden, and increased across remodeling phenotypes. This breadth is consistent with its established role as an integrative marker of wall stress and cumulative myocardial burden ^29–31^ and may explain why natriuretic peptides perform well for risk stratification but are less suited to localizing an individual patient within the remodeling sequence.

By contrast, the matrix turnover markers were domain-selective. TIMP-1 and Galectin-3 were each associated with native T1 and with no other remodeling parameter. In proportional-odds models, TIMP-1 was associated with remodeling phenotype category in both unadjusted and adjusted models (adjusted OR 2.18 per SD, 95% CI 1.37-3.48), and the association strengthened rather than attenuated after adjustment for age, sex, and renal function, arguing against confounding by the principal non-cardiac determinants of both matrix-marker concentrations and native T1. These observations are in line with experimental data implicating TIMP-1 and Galectin-3 in fibroblast activation and extracellular matrix remodeling in the pressure-overloaded myocardium ^18–21^, with the reported correlation of circulating TIMP-1 with histologically quantified fibrosis in severe AS ^32^ and with its association with survival after transcatheter aortic valve replacement ^33^. The present findings extend these observations to moderate AS and to a defined imaging correlate rather than to stenosis severity or clinical outcome.

Notably, the matrix-marker associations were confined to native T1 and did not extend to ECV (β=0.12-0.15). Native T1 and ECV are related but non-interchangeable measures: native T1 reflects a composite of interstitial and intracellular signal without contrast administration, whereas ECV isolates the extracellular fraction, at reduced complete-case n in this cohort and with additional hematocrit-related variability. Whether the matrix-turnover signal is genuinely more closely related to the composite T1 signal, or whether ECV associations were missed at the available precision (NT-proBNP-ECV: β=0.35, q=0.046, indicating that ECV was not uninformative), cannot be resolved here and warrants examination in larger cohorts.

sST2 demonstrated a modest inverse association with LV mass index that reached significance only in the primary covariate model. Given that extracardiac sources may contribute substantially to circulating sST2 in disease states ^34^, this observation should be regarded as hypothesis-generating. Notably, high-sensitivity troponin T was not associated with any remodeling parameter after correction for multiplicity, in contrast to reports from severe AS cohorts in which troponin concentrations parallel hypertrophy and adverse outcome ^16,17^. Two explanations appear plausible. First, the dynamic range of troponin concentrations in the present cohort was restricted (median 20 ng/L), limiting the ability to detect graded associations. Second, troponin release is considered to reflect ongoing cardiomyocyte injury accompanying replacement fibrosis, whereas only 30% of the present cohort was LGE-positive, with low overall burden. Both considerations suggest that troponin may become informative in a later category of the disease course than markers of interstitial matrix turnover.

### Diffuse interstitial fibrosis as the biologically active compartment in moderate AS

Collectively, these findings identify diffuse interstitial fibrosis as the compartment in which measurable biological activity is concentrated in moderate AS, based on the studied biomarkers. Native T1 was the only primary diffuse tissue measure associated with TIMP-1 and Galectin-3. Elevated native T1 is sensitive to diffuse myocardial abnormality but is not specific for collagen expansion, and it may also reflect edema, infiltration, or other compositional changes. ECV is more directly related to extracellular expansion, yet neither matrix-related biomarker was associated with ECV. Interstitial matrix expansion precedes replacement fibrosis and systolic dysfunction, was present in the majority of the present cohort, and, in contrast to replacement fibrosis, has been shown to regress after relief of pressure overload ^9,10^. Consistent with this, multiparametric CMR studies have demonstrated tissue-level reverse remodeling after transcatheter aortic valve replacement, including regression of elevated myocardial T2 values, suggestive for tissue edema ^35^. It was also the only compartment to which the matrix turnover markers mapped selectively. The concordance between an imaging measure of interstitial expansion and circulating markers of the matrix-regulatory machinery that generates it, at a stage preceding established scar, constitutes the central observation of this study. The preservation of effect magnitude after exclusion of patients with infarct-pattern LGE argues against confounding by antecedent ischemic injury, although statistical significance was not retained for all associations in the reduced sample.

### Clinical implications

Recent trials including EARLY TAVR, EVOLVED, and TAVR UNLOAD, reflect growing recognition that myocardial disease burden may rival valve hemodynamics in determining outcome, while illustrating the difficulty of identifying the optimal stage for intervention ^36–38^. A shared feature of these trials is patient selection anchored on markers of advanced myocardial disease, including established midwall LGE, systolic dysfunction, or integrative burden markers such as natriuretic peptides and troponin ^38–40^. If the therapeutic window precedes the establishment of replacement fibrosis, patient selection may require markers indexing earlier interstitial change. The present data support diffuse interstitial fibrosis as such a candidate stage and suggest a pragmatic route toward its identification. Because CMR with T1 mapping cannot feasibly be performed in all patients with moderate AS, circulating markers of matrix turnover that track native T1, like TIMP-1 in the present cohort, may serve as a gatekeeper to advanced tissue characterization rather than a replacement for it. Whether such a biomarker-guided imaging strategy improves selection for earlier aortic valve replacement is a testable hypothesis that requires prospective evaluation in cohorts with serial imaging and, ideally, paired histological or post-AVR regression data.

### Study limitations

Several limitations merit consideration. First, the present analysis is cross-sectional. Causal, compensatory, and parallel-burden interpretations of the biomarker-imaging associations cannot be distinguished, and biomarkers were sampled at a single timepoint, whereas matrix turnover is a dynamic process. Second, circulating markers of matrix turnover, rather than turnover itself, were measured. In the absence of paired matrix-metalloproteinase measurement or myocardial histology, the mechanistic interpretation of the TIMP-1 findings rests on published experimental and histological evidence ^18,20,32^ rather than on the present data. Third, this was a targeted investigation of prespecified candidate markers and should not be interpreted as biomarker discovery. Fourth, phenotype assignment relied on prespecified thresholds derived from local healthy-volunteer reference values, and alternative thresholds would redistribute patients between adjacent stages. Fifth, exploratory clinical follow-up accrued 18 events. No biomarker or imaging parameter was associated with the composite endpoint after correction for multiple comparisons, and the cohort is not powered for prognostic inference, which is accordingly not drawn. Finally, external validation in larger prospective cohorts with longer follow-up is required before clinical application can be considered.

## Data Availability

All data produced in the present study are available upon reasonable request to the authors

## Conclusions

In moderate AS, myocardial remodeling is heterogeneous and dissociated from valve hemodynamic severity, and circulating biomarkers index the myocardial rather than the valvular axis of the disease. Markers of extracellular matrix turnover - TIMP-1 in particular - are selectively associated with diffuse interstitial fibrosis and increase across imaging-defined remodeling phenotypes, identifying interstitial matrix expansion as the biologically active compartment at this stage of AS. These findings support a myocardium-centered characterization of aortic stenosis in which advanced imaging and selected circulating biomarkers provide complementary rather than redundant information.

### Sources of Funding

This work was funded by the Deutsche Forschungsgemeinschaft (DFG, German Research Foundation) – Grant No. 397484323 – TRR259, project C06.

## Supplemental Material

Supplemental Methods, Supplemental Tables S1–S7, Supplemental Figures S1–S4

## Nonstandard Abbreviations and Acronyms

AS: Aortic stenosis
AVR: Aortic valve replacement
CMR: Cardiac magnetic resonance imaging
ECV: Extracellular volume fraction
EDTA: Ethylenediaminetetraacetic acid
Gal-3: Galectin-3
LV: Left ventricle
LVEF: Left ventricular ejection fraction
LGE: Late gadolinium enhancement
iAVA: Indexed aortic valve area
iECV: Indexed extracellular volume
TIMP-1: Tissue inhibitor of metalloproteinase-1
sST2: Soluble suppression of tumorigenicity 2

## Supplemental Material

### Supplemental Methods

#### Study Rationale and Eligibility Criteria

The MODAS study (“Biomarkers and Mechanisms of Disease Progression and Outcome of Aortic Stenosis in Humans”), conducted within collaborative research centre TRR259 (project C06), characterizes patients with moderate aortic stenosis by comprehensive multimodal imaging and biomarker profiling at baseline, with longitudinal follow-up for untreated progression to severe aortic stenosis and aortic valve replacement. Eligible patients for the present analysis had acquired, tricuspid moderate aortic stenosis under outpatient cardiological care and were older than 18 years. The diagnosis of moderate AS was made based on current ESC echocardiographic criteria ^41^. Exclusion criteria were a bicuspid aortic valve, mild or severe aortic stenosis, contraindications to cardiac magnetic resonance or computed tomography, inability to comply with study procedures, and absence of informed consent. Demographics, cardiovascular risk factors, comorbidities and NYHA functional class were recorded by structured interview and electronic health record review.

#### Cardiac Magnetic Resonance Acquisition

CMR examinations were performed in supine position using a commercially available 1.5 T system (Ingenia, Philips Healthcare, Best, The Netherlands) with retrospective ECG-gating and a 28-channel coil for cardiac imaging. Sequences were acquired during end-expiratory breath-hold. The protocol comprised balanced steady-state free precession cine sequences, including a contiguous short-axis stack covering the left ventricle from base through apex, standard 2-, 3-, and 4-chamber long-axis views, and an additional cine plane perpendicular to the aortic valve.

Prior to administration of gadolinium contrast, T1-mapping (using a 5(3)3 Modified Look-Locker inversion recovery (MOLLI) sequence) was acquired in SA (three equally spaced slices: apical, mid-ventricular, and basal). Afterwards, automatic injection of Gadobutrol (Gadovist, Bayer HealthCare Pharmaceuticals, Berlin, Germany, 0.2 mmol/kg) was initiated into an antecubital vein (flow rate of 2 mL/s) followed by a 20 mL saline flush. 10 min post contrast, T1-mapping was acquired in matching locations in SA. For LGE imaging, T1-weighted inversion-recovery fast spoiled gradient-echo sequences were used and acquired in the same orientations as cine sequences with inversion time adjusted to null LV myocardium. Detailed sequence parameters are provided in Supplementary Table S1.

#### CMR Image Analysis

CMR studies were analyzed using commercially available software (MR Cardiac Suite, Philips Advanced Visualization Workspace, release 15.0.7). Two cardiovascular radiologists with 5 (JJ) and 9 (LP) years of experience in CMR analyzed all datasets and performed measurements in consensus based on standardized image interpretation guidelines ^42^. Readers were blinded to biomarker concentrations and clinical outcomes.

SA cine images were used for volumetric assessment of the LV, LVEF, and LV mass with the latter consisting of LV wall mass and LV papillary muscle mass. Values were indexed to body surface area (BSA) on the day of examination using the Mosteller method ^43^. Maximum LV wall thickness was measured perpendicular to the myocardial wall on end-diastolic short-axis cine images, excluding papillary muscles and trabeculations. LV hypertrophy was defined by sex-specific LV mass index thresholds (>84 g/m² in men and >69 g/m² in women) ^42^.

Native and postcontrast T1 maps were visually inspected for motion, misregistration, and susceptibility artifacts. Endocardial and epicardial contours were delineated and myocardial values were assigned according to the 16-segment American Heart Association model.

Segments affected by artifacts or partial-volume contamination were excluded. Areas of focal late gadolinium enhancement were excluded from the assessment of diffuse tissue characteristics. Global native T1 and extracellular volume fraction were calculated as the mean of all valid segmental values, and basal values were calculated as the mean of the six basal segments.

Synthetic hematocrit and synthetic ECV were calculated by the vendor-provided method for 1.5-T MOLLI sequences as described by Fent et al. ^44^ and were based on precontrast and postcontrast myocardial and blood-pool T1 values. In addition, indexed extracellular volume (iECV) was calculated as (ECV fraction × indexed LV mass) / (1.05 g/mL × 100) ^28^. Extracellular matrix volume and cellular volume were calculated as ECV fraction × LV mass / (1.05 g/mL) and (1 − ECV fraction) × LV mass / (1.05 g/mL), respectively.

For assessment of replacement fibrosis, SA LGE images were reviewed for the presence of visually detectable enhancement, defined as areas of relatively increased signal intensity after contrast administration. If present, LGE was semi-automatically quantified using the 6-standard deviation technique after manual delineation of a region of interest in remote myocardium, defined as myocardium without visible enhancement or wall motion abnormality ^42^. Segmented areas were summed to derive total LGE burden, which was expressed in grams and as a percentage of LV myocardial wall mass. Typical subendocardial LGE with or without transmural extension in a coronary vascular territory was classified as infarct-related, all other enhancement patterns were classified as non-infarct LGE.

#### Derivation of Native T1 and ECV Thresholds

Established local institutional reference ranges for native T1 and extracellular volume fraction were used (based on scans of 30 healthy volunteers without known cardiovascular disease and with normal CMR findings). Reference participants were examined on the same 1.5-T system using the identical acquisition and postprocessing protocol as the study cohort. The corresponding upper reference limits were used to define elevated native T1 (>985 ms) and elevated extracellular volume fraction (>29%) in the study cohort.

#### Biomarker Measurement

Peripheral venous blood was drawn at baseline into EDTA tubes, centrifuged at 3000 g for 10 minutes at room temperature within 60 minutes of collection, and the separated plasma stored at −80°C until batch analysis. Samples underwent 1 freeze–thaw cycle before measurement.

Plasma concentrations of soluble suppression of tumorigenicity 2 (sST2), tissue inhibitor of metalloproteinase-1 (TIMP-1), and Galectin-3 (Gal-3) were measured by quantitative solid-phase sandwich enzyme-linked immunosorbent assay (ELISA) using commercially available kits (R&D Systems/Bio-Techne, Minneapolis, Minnesota, catalog numbers DY523B-05 for sST2, DY970 for TIMP-1, and DY1154 for Gal-3) according to the manufacturer’s instructions. Samples were diluted 1:100 (sST2), 1:250 (TIMP-1), and 1:8 (Gal-3) as required to fall within the linear range of each assay, and measured in singlicate. Absorbance was read at 450 nm with wavelength correction at 540 nm. Concentrations were interpolated from a 4-parameter logistic standard curve.

N-terminal pro–B-type natriuretic peptide (NT-proBNP) and high-sensitivity troponin T (hs-TnT) were measured in the accredited institutional clinical chemistry laboratories using standard electrochemiluminescence immunoassays. The hs-TnT assay had a 99th-percentile upper reference limit of 14 ng/L. All laboratory personnel were blinded to imaging findings and clinical outcomes.

## Supplemental Statistical Methods

Univariable associations displayed in the figure panels used Pearson correlation of log-transformed biomarker concentrations with CMR parameters and with echocardiographic severity indices, and Pearson correlation of severity indices with the phenotype-defining CMR parameters. Adjusted linear models entered the standardized log-biomarker after age, sex, and eGFR.

This parsimonious covariate set was prespecified to limit model complexity, relative to the available sample size of 89 patients and 18 events and to avoid risking overfitting. The Benjamini-Hochberg procedure was applied within each prespecified test family (20 confirmatory biomarker-CMR tests, 55 exploratory biomarker-CMR tests, 15 biomarker-severity tests, 12 severity-CMR tests, 5 ordinal phenotype models, and 10 outcome predictors). Proportional-odds ordinal models were fitted by maximum likelihood with Wald inference. Cox proportional-hazards models used the Breslow partial likelihood, with hazard ratios per 1-SD increase for continuous predictors. Kaplan-Meier curves were compared by the log-rank test, and the Gehan-Breslow-Wilcoxon statistic is additionally shown. Odds ratios are maximum-likelihood estimates with Wald 95% confidence intervals. Missing values were treated as missing, and no imputation was performed. Data were frozen on 21 June 2026.

### Sensitivity analyses

The biomarker–CMR and ordinal phenotype analyses were repeated after exclusion of patients with infarct-pattern LGE (Supplemental Table S5). For the exploratory clinical endpoint, native T1 was additionally modeled as a continuous variable and across candidate thresholds from 955 to 1030 ms in 5-ms increments, by logistic and Cox regression.

## Supplemental figures

**Supplemental Figure S1:**
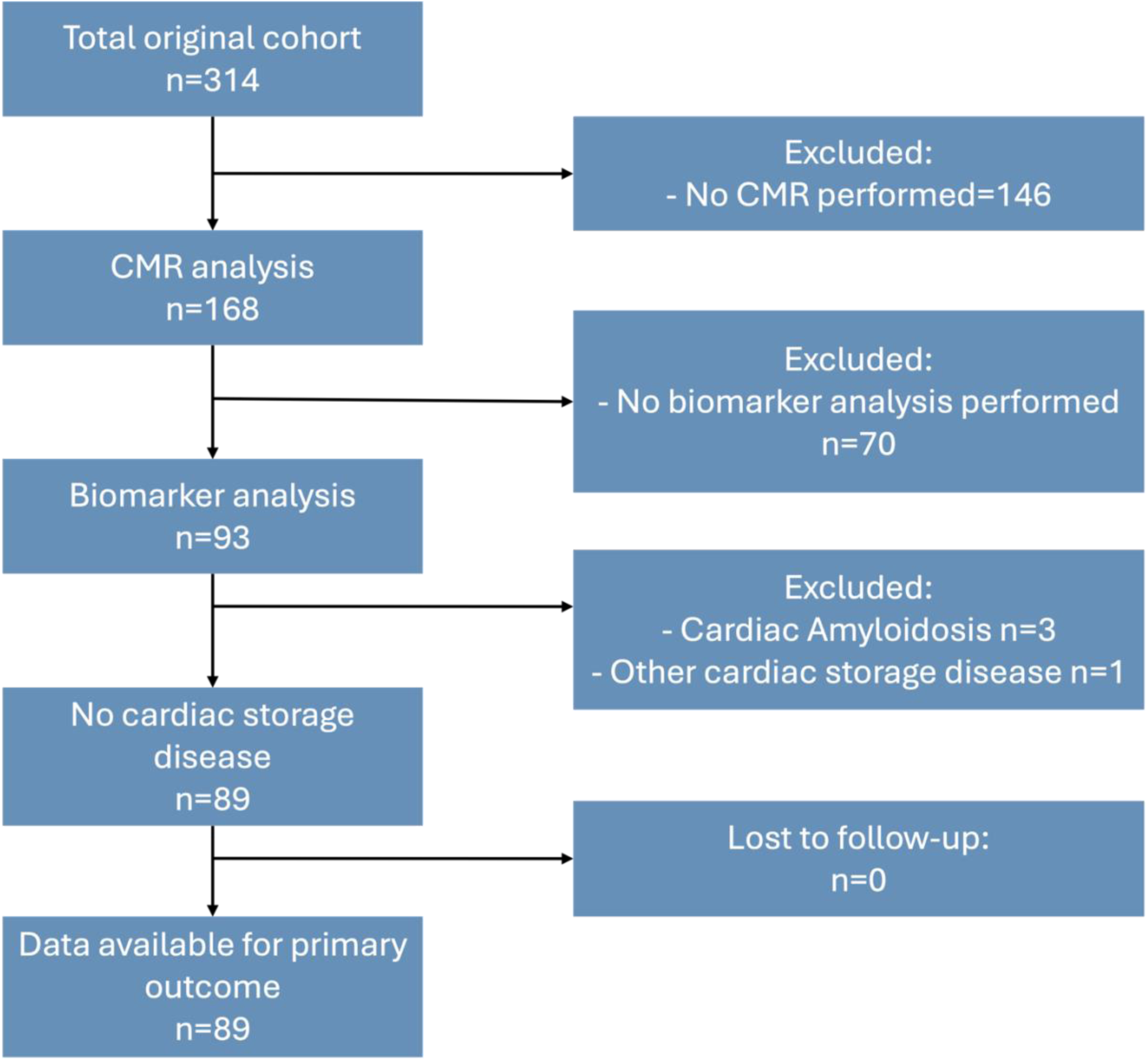
Flow diagram of study participants.

**Supplemental Figure S2.**
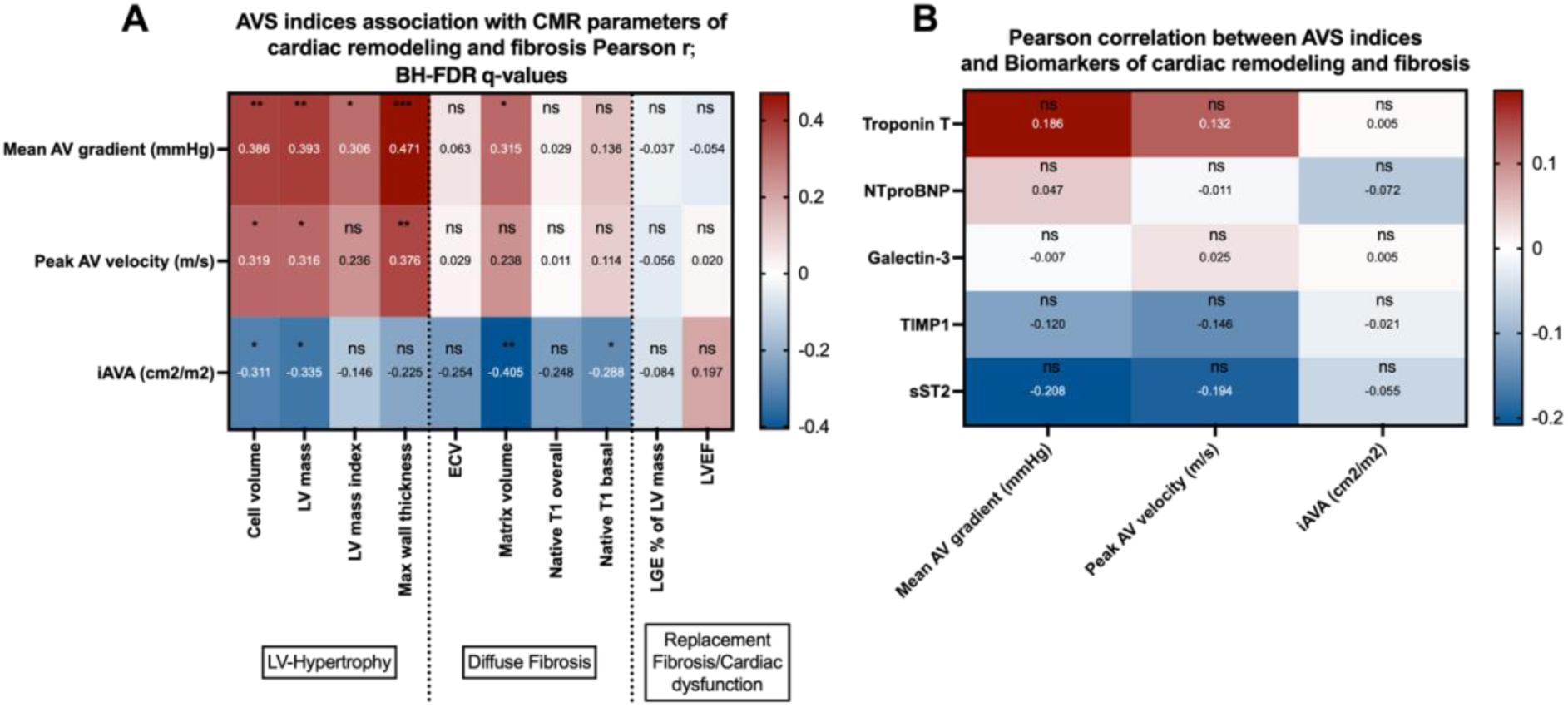
Univariable association of AS severity indices with CMR parameters of cardiac remodeling and fibrosis (A, Pearson, BH-FDR corrected) and of circulating biomarkers with AS severity indices (B, Pearson). ns = not significant, * q<0.05, ** q<0.01, *** q<0.001.

**Supplemental Figure S3.**
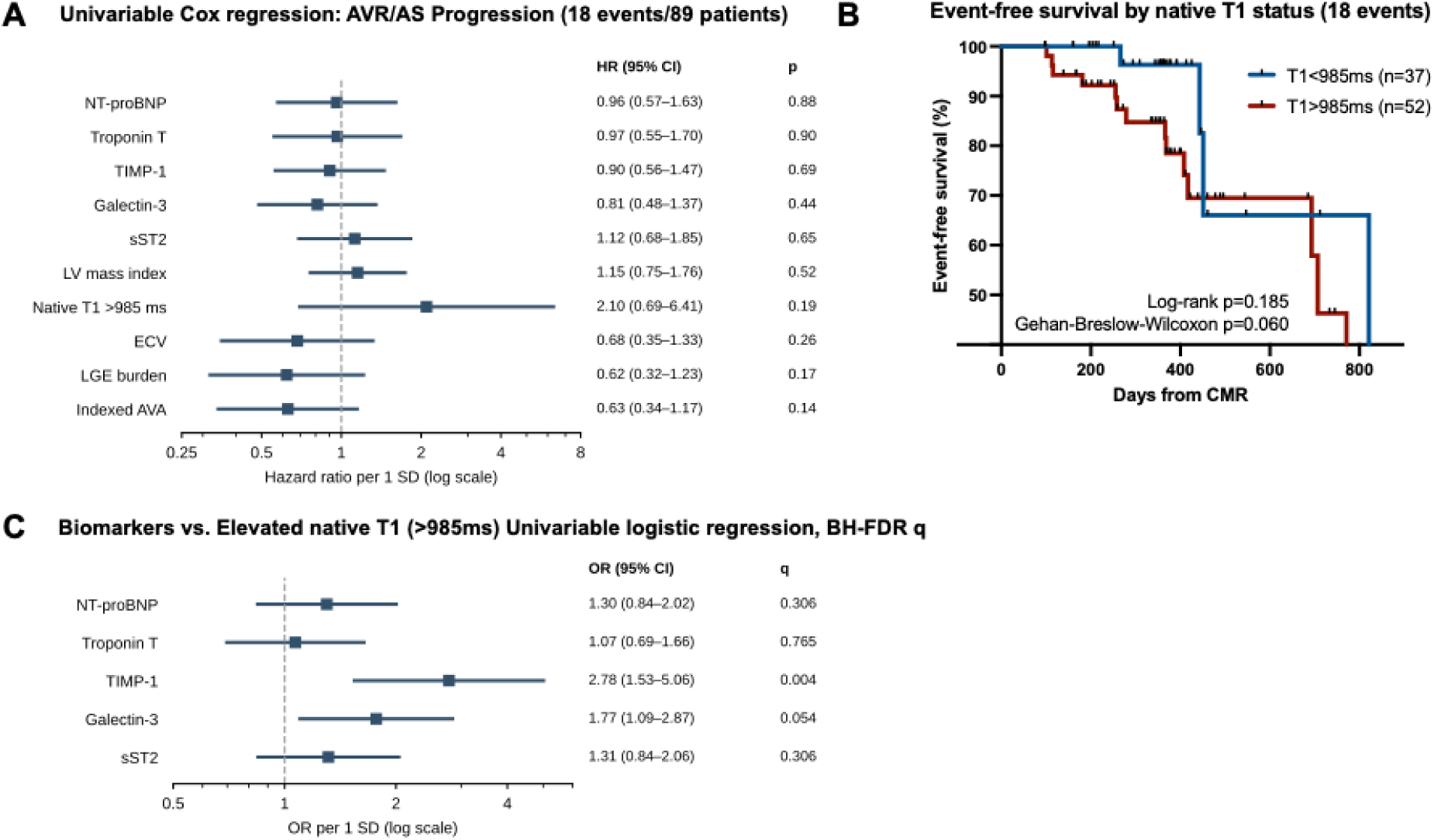
Exploratory clinical follow-up (18 events). No predictor was significantly associated with the endpoint after false-discovery-rate correction (all q≥0.63). No candidate native-T1 threshold between 955 and 1030 ms was significantly associated with the endpoint (all p>0.07). A: Univariable Cox regression, hazard ratios per 1 SD (native T1 as >985 ms vs <=985 ms). B: Kaplan-Meier event-free survival by native T1 status (log-rank and Gehan-Breslow-Wilcoxon tests). C: Univariable logistic regression of elevated native T1 on each biomarker, OR per 1 SD with Benjamini-Hochberg q. No candidate threshold for native T1 between 955 and 1030 ms was significantly associated with the endpoint (all p>0.07)

**Supplemental Figure S4.**
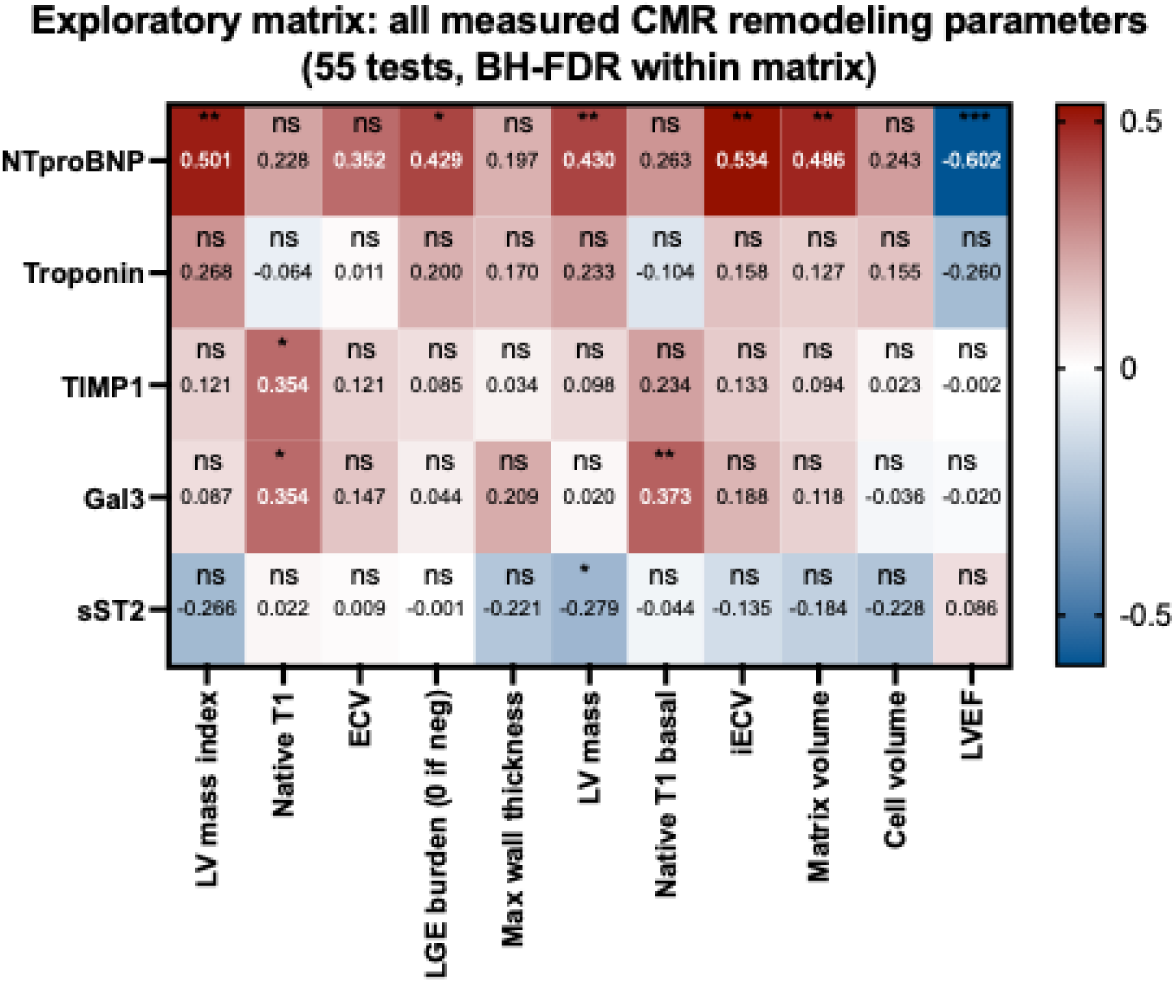
Exploratory matrix: adjusted associations. (standardized β, age-, sex-, eGFR-adjusted) of log-transformed biomarkers with all measured CMR parameters (55 tests, Benjamini-Hochberg q within the matrix). * q<0.05, ** q<0.01, *** q<0.001, ns, not significant.

## Supplemental tables

**Supplemental Table S1:** Cardiovascular magnetic resonance sequence parameters.

|  |  |  |  |
| --- | --- | --- | --- |
|  | Cine | LGE | T1 mapping |
| Sequence | bSSFP | T1w IR fast spoiled GE | 5(3)3 MOLLI |
| Field of view (mm) | 300-350 x 218-350 x 112 | 280-300 x 204-239 x 50 | 340 x 389 x 44 |
| Acquired resolution (mm) | 1.7-1.8 x 1.7-1.9 | 1.64 x 1.72 | 1.98 x 2.0 |
| Reconstructed resolution (mm) | 1.0 x 1.0 | 1.59 x 1.59 | 0.58 x 0.58 |
| Slice thickness (mm) | 8 | 10 | 10 |
| Repetition time (ms) | 3 | 3.5 | 2 |
| Echo time (ms) | 1.5 | 1.7 | 0.8 |
| Flip angle (°) | 60-100 | 15 | 35 |
| Temporal resolution (ms) | ~25 ms* | Ø | Ø |
| Acceleration | Compressed SENSE, factors 4-4.4 | Ø | SENSE, factor 2.2 |
| bSSFP = Balanced steady-state free precession. GE=gradient echo. IR=Inversion recovery. LGE=Late gadolinium enhancement. MOLLI=Modified Look-Locker inversion recovery. SENSE=sensitivity encoding. *=heart rate dependent (40 cardiac phases per R-R interval are acquired, and the provided value refers to a heart rate of 80 beats per minute). |  |  |  |

**Supplemental Table S2:** Patient-level characteristics.

| Supplemental Table S2: Patient-level characteristics |  |  |  |  |
| --- | --- | --- | --- | --- |
|  | Total (n=89) | Stable AS (n=71) | AVR/ AS Progression (n=18) | p-value |
| Body surface area (m2) | 2.00 (1.80-2.10) | 2.00 (1.80-2.16) | 1.90 (1.73-2.00) | 0.0762 |
| **Past medical history** |  |  |  |  |
| Peripheral artery disease | 12 (14.6%) | 9 (14.1%) | 3 (16.7%) | 0.7203 |
| History of/active cancer | 21 (25.3%) | 15 (23.1%) | 6 (33.3%) | 0.3752 |
| Previous stroke | 8 (9.6%) | 7 (10.8%) | 1 (5.6%) | 0.6796 |
| Previous cardiac surgery | 11 (13.3%) | 9 (13.8%) | 2 (11.1%) | 0.9999 |
| Previous/active Smoking | 31 (37.3%) | 27 (41.5%) | 4 (22.2%) | 0.1734 |
| **Laboratory parameters** |  |  |  |  |
| Hemoglobin (g/dL) | 14.0 (12.0-15.0) | 14.0 (12.5-15.0) | 13.0 (12.0-14.8) | 0.2961 |
| White blood cell count (x10<sup>9</sup>/L) | 6.8 (5.3-8.1) | 6.8 (5.3-8.1) | 6.8 (5.9-7.9) | 0.9829 |
| Platelet count (x10<sup>9</sup>/L) | 226 (182-276) | 226 (182-276) | 215 (170-272) | 0.8765 |
| Creatinine (mg/dL) | 1.02 (0.84-1.21) | 1.04 (0.86-1.20) | 1.00 (0.79-1.25) | 0.6238 |
| GFR CKD (ml/min) | 67 (54-81) | 66 (50-84) | 70 (56-76) | 0.6978 |
| LDL-cholesterol (mg/dL) | 76 (60-106) | 76 (62-107) | 76 (56-104) | 0.8907 |
| C-Reactive Protein (mg/L) | 1.9 (0.8-3.8) | 2.0 (0.8-4.2) | 1.2 (0.8-3.6) | 0.4064 |
| Values are median (IQR), mean ± SD, or n (%)<br>AVR=aortic valve replacement, LDL =low-density lipoprotein, GFR=glomerular filtration rate<br>* Progression to severe aortic stenosis AVA <1cm<sup>2</sup>, or mean aortic pressure gradient >40mmHg, or peak aortic velocity >4m/s at follow-up |  |  |  |  |

**Supplemental Table S3:** CMR-based remodeling phenotypes.

| Supplemental Table S3: CMR-based remodeling phenotypes |  |  |  |  |  |
| --- | --- | --- | --- | --- | --- |
|  | Category I<br>No LVH/<br>fibrosis<br>(n=15) | Category II<br>LVH only<br>(n=14) | Category III<br>Diffuse Fibrosis<br>+/- LVH<br>(n=33) | Category IV:<br>LGE positive +/-<br>diffuse fibrosis<br>(n=27) | p-value |
| **Echocardiographic measures at baseline** |  |  |  |  |  |
| Peak aortic valve gradient (mmHg) | 27.5 (21.0-32.9) | 36.0 (29.0-50.0) | 31.9 (23.5-42.5) | 32.0 (24.1-39.5) | 0.2738 |
| Peak aortic valve velocity (m/s) | 2.70 (2.35-2.85) | 3.00 (2.70-3.50) | 2.80 (2.50-3.20) | 2.80 (2.45-3.15) | 0.2501 |
| **Cardiac magnetic resonance parameters of cardiac remodeling and fibrosis** |  |  |  |  |  |
| LV mass (g) | 138.6 (129.8-146.1) | 168.4 (143.0-191.9) | 155.5 (135.9-196.2) | 177.1 (163.5-202.1) | 0.0041 |
| LV mass index (g/m<sup>2</sup>) | 72.4 (65.6-78.2) | 86.2 (80.5-91.4) | 82.9 (70.5-93.8) | 86.0 (81.6-103.4) | 0.0015 |
| Maximum LV wall thickness (mm) | 11.7 (11.2-12.8) | 12.1 (11.4-14.2) | 12.6 (10.7-14.1) | 13.3 (12.1-15.5) | 0.0446 |
| ECV fraction (%) | 23.5 (21.9-25.2) | 25.8 (23.2-27.0) | 27.2 (25.2-29.7) | 27.6 (24.3-30.9) | 0.0007 |
| iECV (mL/m<sup>2</sup>) | 15.8 (13.1-16.5) | 21.5 (18.5-22.9) | 22.9 (19.5-26.2) | 22.6 (19.3-28.0) | <0.0001 |
| Matrix volume (mL) | 29.9 (27.6-34.3) | 40.7 (35.7-45.2) | 43.7 (34.7-52.8) | 43.8 (39.1-57.3) | <0.0001 |
| Cell volume (mL) | 99.6 (96.1-105.7) | 119.3 (103.0-135.9) | 102.3 (90.2-138.3) | 125.3 (111.5-131.5) | 0.0244 |
| Native T1 overall (ms) | 968 (961-975) | 969 (957-978) | 1013 (1004-1034) | 1005 (991-1033) | <0.0001 |
| Native T1 basal (ms) | 974 (962-991) | 980 (959-991) | 1022 (1005-1042) | 1013 (995-1043) | <0.0001 |
| LV ejection fraction (%) | 74 (70-78) | 69 (66-72) | 69 (61-74) | 67 (58-75) | 0.0962 |
| Values are median (IQR), mean ± SD, or n (%) | | | | | |
| AVR=aortic valve replacement. |  |  |  |  |  |
| * Progression to severe aortic stenosis AVA <1cm<sup>2</sup>, or mean aortic pressure gradient >40mmHg, or peak aortic velocity >4m/s at follow-up |  |  |  |  |  |

**Supplemental Table S4.**
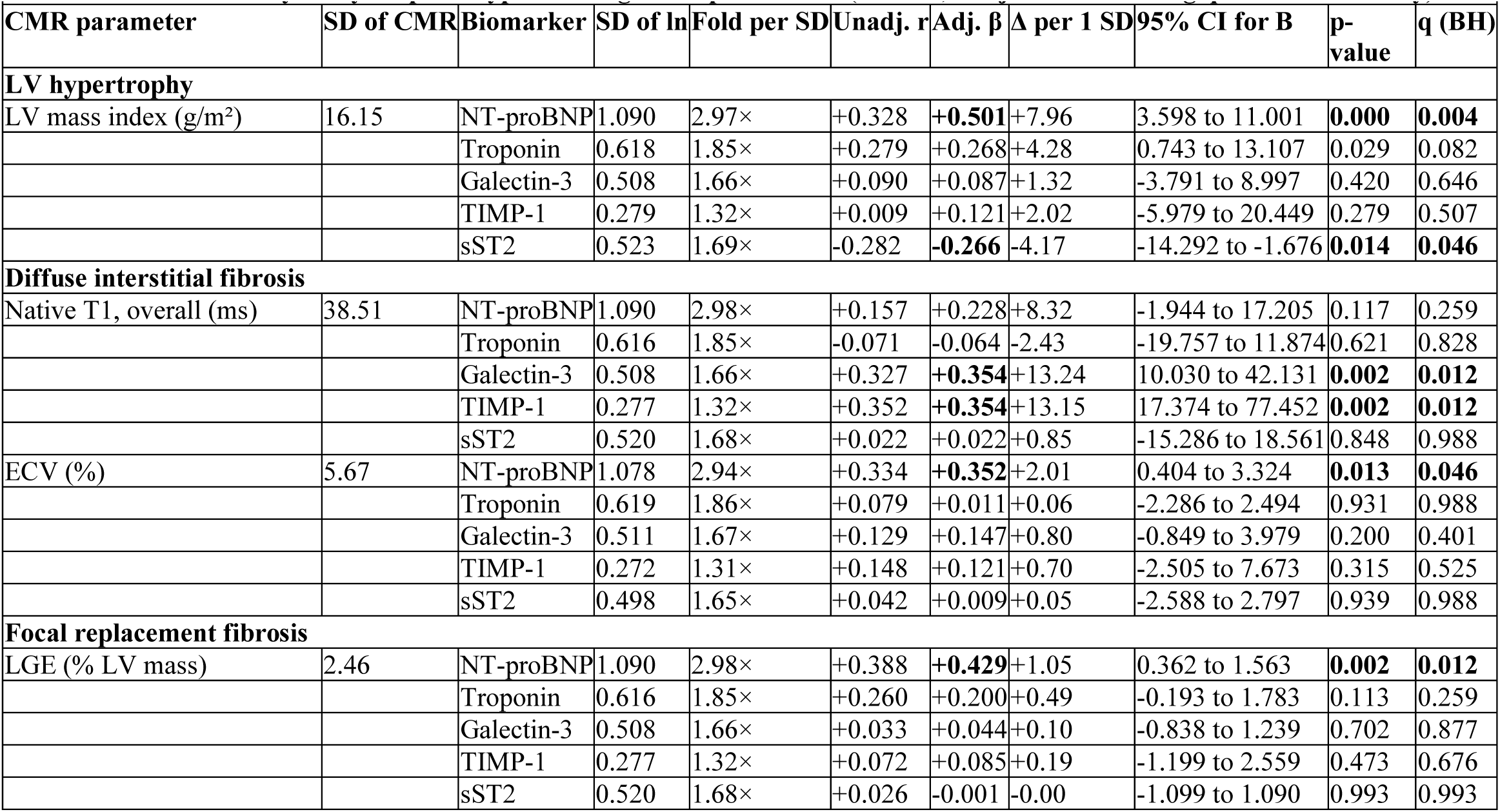
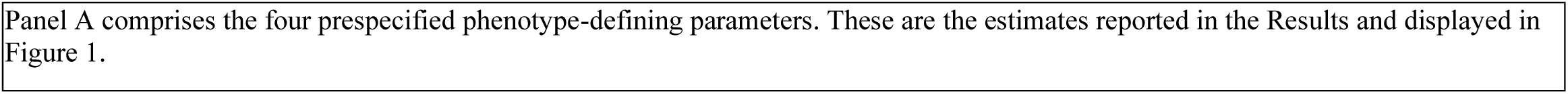
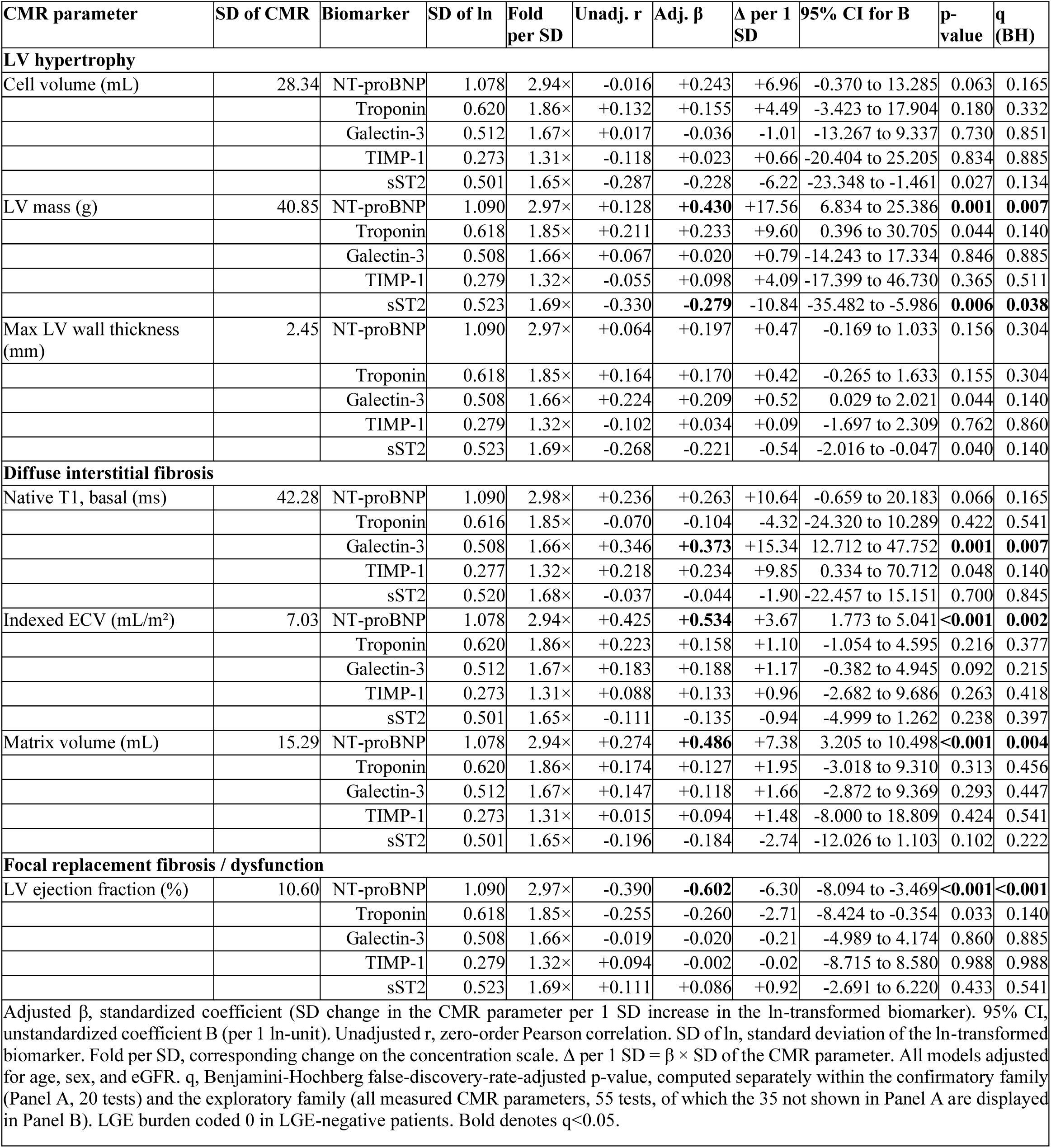
Association between circulating biomarkers and CMR-derived remodeling parameters (multivariable ar regression) panel A — Confirmatory analysis: phenotype-defining CMR parameters (20 tests, Benjamini-Hochberg q within this family) panel B — Exploratory analysis: remaining CMR parameters (35 of the 55 tests of the exploratory family, Benjamini-Hochberg q puted across all 55 tests)

| Supplemental Table S4. Association between circulating biomarkers and CMR-derived remodeling parameters (multivariable linear regression) |  |  |  |  |  |  |  |  |  |  |
| --- | --- | --- | --- | --- | --- | --- | --- | --- | --- | --- |
| Panel A — Confirmatory analysis: phenotype-defining CMR parameters (20 tests, Benjamini-Hochberg q within this family) |  |  |  |  |  |  |  |  |  |  |
| CMR parameter | SD of CMR | Biomarker | SD of ln | Fold per SD | Unadj. r | Adj. $\beta$ | $\Delta$ per 1 SD | 95% CI for B | p-value | q (BH) |
| **LV hypertrophy** |  |  |  |  |  |  |  |  |  |  |
| LV mass index (g/m<sup>2</sup>) | 16.15 | NT-proBNP | 1.090 | 2.97× | +0.328 | **+0.501** | +7.96 | 3.598 to 11.001 | **0.000** | **0.004** |
|  |  | Troponin | 0.618 | 1.85× | +0.279 | +0.268 | +4.28 | 0.743 to 13.107 | 0.029 | 0.082 |
|  |  | Galectin-3 | 0.508 | 1.66× | +0.090 | +0.087 | +1.32 | -3.791 to 8.997 | 0.420 | 0.646 |
|  |  | TIMP-1 | 0.279 | 1.32× | +0.009 | +0.121 | +2.02 | -5.979 to 20.449 | 0.279 | 0.507 |
|  |  | sST2 | 0.523 | 1.69× | -0.282 | **-0.266** | -4.17 | -14.292 to -1.676 | **0.014** | **0.046** |
| **Diffuse interstitial fibrosis** |  |  |  |  |  |  |  |  |  |  |
| Native T1, overall (ms) | 38.51 | NT-proBNP | 1.090 | 2.98× | +0.157 | +0.228 | +8.32 | -1.944 to 17.205 | 0.117 | 0.259 |
|  |  | Troponin | 0.616 | 1.85× | -0.071 | -0.064 | -2.43 | -19.757 to 11.874 | 0.621 | 0.828 |
|  |  | Galectin-3 | 0.508 | 1.66× | +0.327 | **+0.354** | +13.24 | 10.030 to 42.131 | **0.002** | **0.012** |
|  |  | TIMP-1 | 0.277 | 1.32× | +0.352 | **+0.354** | +13.15 | 17.374 to 77.452 | **0.002** | **0.012** |
|  |  | sST2 | 0.520 | 1.68× | +0.022 | +0.022 | +0.85 | -15.286 to 18.561 | 0.848 | 0.988 |
| ECV (%) | 5.67 | NT-proBNP | 1.078 | 2.94× | +0.334 | **+0.352** | +2.01 | 0.404 to 3.324 | **0.013** | **0.046** |
|  |  | Troponin | 0.619 | 1.86× | +0.079 | +0.011 | +0.06 | -2.286 to 2.494 | 0.931 | 0.988 |
|  |  | Galectin-3 | 0.511 | 1.67× | +0.129 | +0.147 | +0.80 | -0.849 to 3.979 | 0.200 | 0.401 |
|  |  | TIMP-1 | 0.272 | 1.31× | +0.148 | +0.121 | +0.70 | -2.505 to 7.673 | 0.315 | 0.525 |
|  |  | sST2 | 0.498 | 1.65× | +0.042 | +0.009 | +0.05 | -2.588 to 2.797 | 0.939 | 0.988 |
| **Focal replacement fibrosis** |  |  |  |  |  |  |  |  |  |  |
| LGE (% LV mass) | 2.46 | NT-proBNP | 1.090 | 2.98× | +0.388 | **+0.429** | +1.05 | 0.362 to 1.563 | **0.002** | **0.012** |
|  |  | Troponin | 0.616 | 1.85× | +0.260 | +0.200 | +0.49 | -0.193 to 1.783 | 0.113 | 0.259 |
|  |  | Galectin-3 | 0.508 | 1.66× | +0.033 | +0.044 | +0.10 | -0.838 to 1.239 | 0.702 | 0.877 |
|  |  | TIMP-1 | 0.277 | 1.32× | +0.072 | +0.085 | +0.19 | -1.199 to 2.559 | 0.473 | 0.676 |
|  |  | sST2 | 0.520 | 1.68× | +0.026 | -0.001 | -0.00 | -1.099 to 1.090 | 0.993 | 0.993 |

| CMR parameter | SD of CMR | Biomarker | SD of ln | Fold per SD | Unadj. r | Adj. $\beta$ | $\Delta$ per 1 SD | 95% CI for B | p-value | q (BH) |
| --- | --- | --- | --- | --- | --- | --- | --- | --- | --- | --- |
| **LV hypertrophy** |  |  |  |  |  |  |  |  |  |  |
| Cell volume (mL) | 28.34 | NT-proBNP | 1.078 | 2.94× | -0.016 | +0.243 | +6.96 | -0.370 to 13.285 | 0.063 | 0.165 |
|  |  | Troponin | 0.620 | 1.86× | +0.132 | +0.155 | +4.49 | -3.423 to 17.904 | 0.180 | 0.332 |
|  |  | Galectin-3 | 0.512 | 1.67× | +0.017 | -0.036 | -1.01 | -13.267 to 9.337 | 0.730 | 0.851 |
|  |  | TIMP-1 | 0.273 | 1.31× | -0.118 | +0.023 | +0.66 | -20.404 to 25.205 | 0.834 | 0.885 |
|  |  | sST2 | 0.501 | 1.65× | -0.287 | -0.228 | -6.22 | -23.348 to -1.461 | 0.027 | 0.134 |
| LV mass (g) | 40.85 | NT-proBNP | 1.090 | 2.97× | +0.128 | **+0.430** | +17.56 | 6.834 to 25.386 | **0.001** | **0.007** |
|  |  | Troponin | 0.618 | 1.85× | +0.211 | +0.233 | +9.60 | 0.396 to 30.705 | 0.044 | 0.140 |
|  |  | Galectin-3 | 0.508 | 1.66× | +0.067 | +0.020 | +0.79 | -14.243 to 17.334 | 0.846 | 0.885 |
|  |  | TIMP-1 | 0.279 | 1.32× | -0.055 | +0.098 | +4.09 | -17.399 to 46.730 | 0.365 | 0.511 |
|  |  | sST2 | 0.523 | 1.69× | -0.330 | **-0.279** | -10.84 | -35.482 to -5.986 | **0.006** | **0.038** |
| Max LV wall thickness (mm) | 2.45 | NT-proBNP | 1.090 | 2.97× | +0.064 | +0.197 | +0.47 | -0.169 to 1.033 | 0.156 | 0.304 |
|  |  | Troponin | 0.618 | 1.85× | +0.164 | +0.170 | +0.42 | -0.265 to 1.633 | 0.155 | 0.304 |
|  |  | Galectin-3 | 0.508 | 1.66× | +0.224 | +0.209 | +0.52 | 0.029 to 2.021 | 0.044 | 0.140 |
|  |  | TIMP-1 | 0.279 | 1.32× | -0.102 | +0.034 | +0.09 | -1.697 to 2.309 | 0.762 | 0.860 |
|  |  | sST2 | 0.523 | 1.69× | -0.268 | -0.221 | -0.54 | -2.016 to -0.047 | 0.040 | 0.140 |
| **Diffuse interstitial fibrosis** |  |  |  |  |  |  |  |  |  |  |
| Native T1, basal (ms) | 42.28 | NT-proBNP | 1.090 | 2.98× | +0.236 | +0.263 | +10.64 | -0.659 to 20.183 | 0.066 | 0.165 |
|  |  | Troponin | 0.616 | 1.85× | -0.070 | -0.104 | -4.32 | -24.320 to 10.289 | 0.422 | 0.541 |
|  |  | Galectin-3 | 0.508 | 1.66× | +0.346 | **+0.373** | +15.34 | 12.712 to 47.752 | **0.001** | **0.007** |
|  |  | TIMP-1 | 0.277 | 1.32× | +0.218 | +0.234 | +9.85 | 0.334 to 70.712 | 0.048 | 0.140 |
|  |  | sST2 | 0.520 | 1.68× | -0.037 | -0.044 | -1.90 | -22.457 to 15.151 | 0.700 | 0.845 |
| Indexed ECV (mL/m<sup>2</sup>) | 7.03 | NT-proBNP | 1.078 | 2.94× | +0.425 | **+0.534** | +3.67 | 1.773 to 5.041 | **<0.001** | **0.002** |
|  |  | Troponin | 0.620 | 1.86× | +0.223 | +0.158 | +1.10 | -1.054 to 4.595 | 0.216 | 0.377 |
|  |  | Galectin-3 | 0.512 | 1.67× | +0.183 | +0.188 | +1.17 | -0.382 to 4.945 | 0.092 | 0.215 |
|  |  | TIMP-1 | 0.273 | 1.31× | +0.088 | +0.133 | +0.96 | -2.682 to 9.686 | 0.263 | 0.418 |
|  |  | sST2 | 0.501 | 1.65× | -0.111 | -0.135 | -0.94 | -4.999 to 1.262 | 0.238 | 0.397 |
| Matrix volume (mL) | 15.29 | NT-proBNP | 1.078 | 2.94× | +0.274 | **+0.486** | +7.38 | 3.205 to 10.498 | **<0.001** | **0.004** |
|  |  | Troponin | 0.620 | 1.86× | +0.174 | +0.127 | +1.95 | -3.018 to 9.310 | 0.313 | 0.456 |
|  |  | Galectin-3 | 0.512 | 1.67× | +0.147 | +0.118 | +1.66 | -2.872 to 9.369 | 0.293 | 0.447 |
|  |  | TIMP-1 | 0.273 | 1.31× | +0.015 | +0.094 | +1.48 | -8.000 to 18.809 | 0.424 | 0.541 |
|  |  | sST2 | 0.501 | 1.65× | -0.196 | -0.184 | -2.74 | -12.026 to 1.103 | 0.102 | 0.222 |
| **Focal replacement fibrosis / dysfunction** |  |  |  |  |  |  |  |  |  |  |
| LV ejection fraction (%) | 10.60 | NT-proBNP | 1.090 | 2.97× | -0.390 | **-0.602** | -6.30 | -8.094 to -3.469 | **<0.001** | **<0.001** |
|  |  | Troponin | 0.618 | 1.85× | -0.255 | -0.260 | -2.71 | -8.424 to -0.354 | 0.033 | 0.140 |
|  |  | Galectin-3 | 0.508 | 1.66× | -0.019 | -0.020 | -0.21 | -4.989 to 4.174 | 0.860 | 0.885 |
|  |  | TIMP-1 | 0.279 | 1.32× | +0.094 | -0.002 | -0.02 | -8.715 to 8.580 | 0.988 | 0.988 |
|  |  | sST2 | 0.523 | 1.69× | +0.111 | +0.086 | +0.92 | -2.691 to 6.220 | 0.433 | 0.541 |

Adjusted $\beta$ , standardized coefficient (SD change in the CMR parameter per 1 SD increase in the ln-transformed biomarker). 95% CI, unstandardized coefficient B (per 1 ln-unit). Unadjusted r, zero-order Pearson correlation. SD of ln, standard deviation of the ln-transformed biomarker. Fold per SD, corresponding change on the concentration scale. $\Delta$ per 1 SD = $\beta \times$ SD of the CMR parameter. All models adjusted for age, sex, and eGFR. q, Benjamini-Hochberg false-discovery-rate-adjusted p-value, computed separately within the confirmatory family (Panel A, 20 tests) and the exploratory family (all measured CMR parameters, 55 tests, of which the 35 not shown in Panel A are displayed in Panel B). LGE burden coded 0 in LGE-negative patients. Bold denotes $q < 0.05$ .

**Supplemental Table S5.** Biomarker-CMR associations after exclusion of patients with infarct pattern LGE (n=77 Confirmatory analysis in patients without infarct-pattern LGE (20 tests, Benjamini-Hochberg q within this family)

|  |  |  |  |  |  |  |  |  |  |  |
| --- | --- | --- | --- | --- | --- | --- | --- | --- | --- | --- |
| CMR parameter | SD of<br>CMR | Biomarker | SD of<br>ln | Fold<br>per SD | Unadj.<br>r | Adj. $\beta$ | $\Delta$ per 1<br>SD | 95% CI<br>for B | p-value | q (BH) |
| **LV hypertrophy** |  |  |  |  |  |  |  |  |  |  |
| LV mass index (g/m<sup>2</sup>) | 16.04 | NT-proBNP | 1.058 | 2.88× | +0.318 | **+0.528** | +8.43 | 3.825 to<br>12.120 | **<0.001** | **0.006** |
|  |  | Troponin | 0.615 | 1.85× | +0.214 | +0.203 | +3.27 | -1.441 to<br>12.077 | 0.121 | 0.338 |
|  |  | Galectin-3 | 0.511 | 1.67× | +0.078 | +0.060 | +0.90 | -5.239 to<br>8.742 | 0.618 | 0.773 |
|  |  | TIMP-1 | 0.275 | 1.32× | +0.028 | +0.163 | +2.69 | -5.262 to<br>24.783 | 0.199 | 0.442 |
|  |  | sST2 | 0.548 | 1.73× | -0.319 | -0.309 | -4.77 | -15.122 to<br>-2.295 | 0.009 | 0.050 |
| **Diffuse interstitial fibrosis** |  |  |  |  |  |  |  |  |  |  |
| Native T1, overall (ms) | 37.19 | NT-proBNP | 1.060 | 2.89× | +0.247 | +0.316 | +11.64 | 0.474 to<br>21.490 | 0.041 | 0.136 |
|  |  | Troponin | 0.614 | 1.85× | -0.026 | -0.022 | -0.85 | -18.177 to<br>15.402 | 0.870 | 0.902 |
|  |  | Galectin-3 | 0.511 | 1.67× | +0.319 | **+0.365** | +13.71 | 9.751 to<br>43.964 | **0.003** | **0.026** |
|  |  | TIMP-1 | 0.274 | 1.31× | +0.326 | +0.334 | +12.22 | 11.007 to<br>78.359 | 0.010 | 0.050 |
|  |  | sST2 | 0.544 | 1.72× | +0.031 | +0.025 | +0.92 | -15.036 to<br>18.419 | 0.841 | 0.902 |
| ECV (%) | 5.00 | NT-proBNP | 1.048 | 2.85× | +0.338 | +0.349 | +1.78 | 0.258 to<br>3.146 | 0.022 | 0.086 |
|  |  | Troponin | 0.617 | 1.85× | +0.134 | +0.082 | +0.42 | -1.560 to<br>2.922 | 0.546 | 0.728 |
|  |  | Galectin-3 | 0.515 | 1.67× | +0.101 | +0.120 | +0.56 | -1.164 to<br>3.336 | 0.338 | 0.523 |
|  |  | TIMP-1 | 0.268 | 1.31× | +0.143 | +0.132 | +0.68 | -2.493 to<br>7.543 | 0.319 | 0.523 |
|  |  | sST2 | 0.520 | 1.68× | -0.070 | -0.103 | -0.53 | -3.481 to<br>1.436 | 0.409 | 0.584 |
| **Focal replacement fibrosis** |  |  |  |  |  |  |  |  |  |  |
| LGE (% LV mass) | 1.34 | NT-proBNP | 1.060 | 2.89× | +0.311 | +0.226 | +0.31 | -0.094 to<br>0.684 | 0.135 | 0.338 |
|  |  | Troponin | 0.614 | 1.85× | +0.135 | +0.051 | +0.07 | -0.475 to<br>0.702 | 0.702 | 0.826 |
|  |  | Galectin-3 | 0.511 | 1.67× | +0.107 | +0.143 | +0.19 | -0.259 to<br>1.004 | 0.243 | 0.485 |
|  |  | TIMP-1 | 0.274 | 1.31× | +0.081 | +0.016 | +0.02 | -1.235 to<br>1.399 | 0.902 | 0.902 |
|  |  | sST2 | 0.544 | 1.72× | -0.082 | -0.116 | -0.16 | -0.885 to<br>0.310 | 0.340 | 0.523 |
| Layout, model specification, and column definitions as in Supplemental Table S4. Analyses restricted to the 77 patients without infarct-pattern late gadolinium enhancement. All models adjusted for age, sex, and eGFR. q, Benjamini-Hochberg false-discovery-rate-adjusted p-value across the 20 tests. Bold denotes q<0.05. |  |  |  |  |  |  |  |  |  |  |

**Supplemental Table S6.** Patient-level characteristics stratified by the exploratory composite endpoint.

|  |  |  |  |
| --- | --- | --- | --- |
|  | **Stable AS (n=71)** | **AVR/AS progression (n=18)** | **p-value** |
| Age (years) | 75.8 (69.2-81.9) | 76.8 (71.2-79.7) | 0.6715 |
| Women | 19 (26.8%) | 3 (16.7%) | 0.5435 |
| Body mass index (kg/m<sup>2</sup>) | 28.0 (24.0-31.0) | 26.0 (23.2-28.8) | 0.2666 |
| NYHA functional class |  |  | 0.8039 |
| I | 26 (36.6%) | 7 (38.9%) |  |
| II | 25 (35.2%) | 8 (44.4%) |  |
| III | 11 (15.5%) | 2 (11.1%) |  |
| EuroSCORE II (%) | 1.51 (0.87-2.95) | 1.66 (1.40-2.17) | 0.4256 |
| Arterial hypertension | 52 (80.0%) | 15 (83.3%) | 1.0 |
| Hyperlipidemia | 43 (66.2%) | 6 (33.3%) | 0.0159 |
| Diabetes mellitus | 21 (32.3%) | 8 (44.4%) | 0.4059 |
| Coronary artery disease | 30 (46.9%) | 7 (38.9%) | 0.6011 |
| Previous myocardial infarction | 6 (9.2%) | 6 (35.3%) | 0.0145 |
| Atrial fibrillation | 22 (33.8%) | 5 (29.4%) | 1.0 |
| COPD | 7 (10.8%) | 3 (16.7%) | 0.4459 |
| Troponin T (ng/L) | 20.0 (10.0-30.0) | 19.0 (10.0-20.0) | 0.4797 |
| NT-proBNP (pg/mL) | 320 (147-778) | 362 (144-647) | 0.8778 |
| Galectin-3 (pg/mL) | 5103 (3210-6477) | 3989 (3009-6360) | 0.4746 |
| TIMP-1 (pg/mL) | 101255 (85496-121723) | 103871 (95071-121592) | 0.4706 |
| sST2 (pg/mL) | 21373 (15405-27422) | 23762 (18469-26995) | 0.4829 |
| Values are median (IQR), mean ± SD, or n (%) | | | |
| AVR=aortic valve replacement, NYHA=New York Heart Association, COPD=chronic obstructive pulmonary disease |  |  |  |
| * Progression to severe aortic stenosis AVA <1cm<sup>2</sup>, or mean aortic pressure gradient >40mmHg, or peak aortic velocity >4m/s at follow-up |  |  |  |

**Supplemental Table S7.** Imaging characteristics and clinical outcome stratified by the exploratory composite endpoint.

|  | Stable AS (n=71) | AVR/AS progression* (n=18) | p-value |
| --- | --- | --- | --- |
| Peak AV gradient (mmHg) | 31.8 (24.0-37.3) | 33.9 (26.8-41.2) | 0.3897 |
| Mean AV gradient (mmHg) | 16.9 (12.1-22.0) | 18.8 (14.0-22.0) | 0.3568 |
| Peak AV velocity (m/s) | 2.80 (2.42-3.08) | 2.90 (2.62-3.18) | 0.4216 |
| Indexed AVA (cm<sup>2</sup>/m<sup>2</sup>) | 0.70 (0.63-0.79) | 0.59 (0.55-0.71) | 0.0726 |
| Moderate/severe mitral regurgitation | 7 (10.1%) | 3 (16.7%) | 0.4253 |
| Moderate/severe tricuspid regurgitation | 9 (13.0%) | 5 (29.4%) | 0.1395 |
| Cell volume (mL) | 108.9 (97.1-130.0) | 119.4 (100.0-135.4) | 0.5493 |
| LV mass (g) | 157.6 (137.1-184.2) | 167.2 (140.3-195.7) | 0.6122 |
| LV mass index (g/m<sup>2</sup>) | 82.2 (73.1-93.2) | 84.4 (80.6-103.6) | 0.2362 |
| Max LV wall thickness (mm) | 12.6 (11.4-14.1) | 12.6 (11.7-14.1) | 0.9258 |
| ECV fraction (%) | 26.4 (24.2-28.5) | 25.1 (22.9-29.2) | 0.4609 |
| iECV (mL/m<sup>2</sup>) | 21.1 (16.8-24.9) | 20.5 (19.1-26.7) | 0.6803 |
| Native T1 overall (ms) | 991 (972-1019) | 1008 (987-1027) | 0.4227 |
| Native T1 basal (ms) | 1002 (980-1030) | 1015 (996-1043) | 0.2722 |
| Native T1 >985 ms | 38 (53.5%) | 14 (77.8%) | 0.1064 |
| LGE positive | 22 (31.0%) | 5 (27.8%) | 1.0 |
| Infarct-pattern LGE | 10 (14.1%) | 2 (11.1%) | 1.0 |
| Non-infarct LGE | 12 (16.9%) | 3 (16.7%) | 1.0 |
| LGE (% of LV mass, 0 if LGE-negative) | 0.0 (0.0-1.5) | 0.0 (0.0-0.6) | 0.6879 |
| LVEF (%) | 70 (62-75) | 68 (56-72) | 0.2217 |
| Follow-up (days) | 361 (248-412) | 367 (256-449) | 0.7207 |
| Death (any, descriptive) | 1 (1.4%) | 0 (0.0%) | 1.0 |
| Composite events (AVR or severe-AS progression) | 0 | 18 (100%) |  |

Values are median (IQR), mean ± SD, or n (%)
AV=aortic valve, AVR=aortic valve replacement. \* Progression to severe aortic stenosis AVA <1cm<sup>2</sup>, or mean aortic pressure gradient >40mmHg, or peak aortic velocity >4m/s at follow-up

